# The potential impact of novel tuberculosis vaccines on health equity and financial protection in low- and middle-income countries

**DOI:** 10.1101/2022.10.29.22281678

**Authors:** Allison Portnoy, Rebecca A. Clark, Chathika K. Weerasuriya, Christinah Mukandavire, Matthew Quaife, Roel Bakker, Inés Garcia Baena, Nebiat Gebreselassie, Matteo Zignol, Mark Jit, Richard G. White, Nicolas A. Menzies

**Author notes:** **Corresponding Author:** Allison Portnoy, Center for Health Decision Science, Harvard T.H. Chan School of Public Health, 718 Huntington Avenue 2nd Floor, Boston MA 02115, USA.

## Abstract

**Background:** One in two patients developing tuberculosis (TB) in low- and middle-income countries (LMICs) faces catastrophic household costs. We assessed the potential financial risk protection from introducing novel TB vaccines, and how health and economic benefits would be distributed across income quintiles.

**Methods:** We modelled the impact of introducing TB vaccines meeting the WHO preferred product characteristics in 105 LMICs. For each country, we assessed the distribution of health gains, patient costs, and household financial vulnerability following introduction of an infant vaccine, and separately for an adolescent/adult vaccine, compared to a ‘no-new-vaccine’ counterfactual. Patient-incurred direct and indirect costs of TB disease exceeding 20% of annual household income were defined as catastrophic.

**Findings:** Over 2028–2050, the health gains resulting from vaccine introduction were greatest in lower income quintiles, with the poorest two quintiles in each country accounting for 56% of total LMIC TB cases averted. Over this period, the infant vaccine was estimated to avert $5·9 (95% uncertainty interval: $5·3–6·5) billion in patient-incurred total costs, and the adolescent/adult vaccine was estimated to avert $38·9 ($36·6–41·5) billion. Over this period, 3·7 (3·3–4·1) million fewer households were projected to face catastrophic costs with the infant vaccine, and 22·9 (21·4–24·5) million with the adolescent/adult vaccine, with 66% of these gains accruing in the poorest two income quintiles.

**Interpretation:** Under a range of assumptions, introducing novel TB vaccines would reduce income-based inequalities in the health and household economic outcomes of TB in LMICs.

**Funding:** World Health Organization (2020/985800-0)

**Research in context:** *Evidence before this study:* We searched MEDLINE using the terms “tuberculosis,” “catastrophic cost*,” and “vaccin*” and failed to find any original research articles that estimated the impact of TB vaccination on catastrophic costs incurred by TB patients. Previous studies have shown that improved TB prevention and care can lower patient costs and reduce the number of TB-affected households experiencing catastrophic costs, and previous modelling has estimated the potential impact of TB vaccination on patient costs in country case-studies. Survey evidence from high-burden countries have consistently demonstrated higher disease burden among poorer individuals, with TB prevalence in the lowest income quintile on average 2·3 times greater than estimated for the highest income quintile. TB patient cost surveys in high-burden countries have shown that TB patients experience high out-of-pocket and indirect costs, and that these costs represent a greater share of annual household income in poorer income quintiles.

*Added value of this study:* This is the first study to examine the potential for new vaccines to reduce the number of households experiencing catastrophic costs due to TB, and how both these benefits and health gains are distributed across income quintiles. Across all modelled countries over 2028–2050, an adolescent/adult vaccine was projected to reduce TB incidence in the poorest quintile by 13·3 (10·9–15·8) million (30% of total TB cases averted) and reduce the number of households experiencing catastrophic costs by 9·2 (7·5–11·0) million in the poorest quintile (40% of total cases of catastrophic costs averted) compared to the no-new-vaccination baseline.

*Implications of all the available evidence:* Under a range of assumptions, new TB vaccines could be highly impactful and help narrow income-based disparities in the health and the economic consequences of TB for low- and middle-income countries.

## Background

In 2021, 1·5 million individuals died of tuberculosis (TB).^1^ For individuals surviving TB, the disease episode represents an extended period of ill-health, which may lead to chronic disability.^2,3^ This burden of disease is not evenly distributed across the population, with many TB risk factors—crowded living conditions, malnutrition, HIV, and other factors that impair immune function—concentrated in poor and marginalized communities.^4^ Limited healthcare access in these communities also means that individuals developing TB may not receive prompt treatment, extending the duration and severity of disease. Nationally-representative TB prevalence surveys conducted in high-burden countries have consistently demonstrated higher disease burden among poorer individuals, with TB prevalence in the lowest income quintile on average 2·3 times greater than estimated for the highest income quintile.^5-7^

In addition to the individual health effects, TB can have major economic consequences, especially for poor households.^8^ Individuals sick with TB may be less able to work, resulting in income losses. TB-associated healthcare typically involves substantial out-of-pocket costs for patients, despite government-provided TB treatment being free in many countries. Observational studies have shown that individuals with TB frequently make several care-seeking attempts before an accurate diagnosis is made,^9^ which involves additional costs. For poorer households, these costs can represent a substantial share of available income, increasing the risks of facing catastrophic costs.^8^ National survey evidence shows that one in two TB-affected households face costs exceeding 20% of household annual pre-disease income or expenditure.^10^

Several new TB vaccine candidates are in late-stage trials, and their successful development could create new opportunities to prevent TB. However, the concentration of TB burden among poor people in low- and middle-income countries has created additional challenges in communicating the value of vaccines, and likely delayed vaccine development. Previous studies have shown that strengthened TB services can lower patient costs and reduce the number of TB-affected households experiencing catastrophic costs.^11,12^ In this study we examined the potential for new vaccines to reduce the economic burden of TB on affected households, and impact health inequalities. To undertake this study, we simulated the impact of vaccine products meeting the WHO preferred product characteristics for a new TB vaccine.^13^ Comparing these vaccination scenarios to a ‘no-new-vaccine’ baseline, we calculated the potential impact on health outcomes, patient-incurred direct and indirect costs, and TB-affected households experiencing catastrophic costs in 105 low- and middle-income countries over the period 2028– 2050. We report how these benefits would be distributed across income quintiles to assess the potential for new TB vaccines to affect income-based inequalities in the health and economic burden of TB.

## Methods

### Vaccination scenarios

We evaluated an infant ‘pre-infection’ vaccine (i.e., efficacious for individuals uninfected at time of vaccination) with 80% efficacy targeting neonates, and an adolescent/adult ‘pre- and post-infection’ vaccine (i.e., efficacious in all individuals without TB disease at time of vaccination) with 50% efficacy, based on WHO preferred product characteristics (PPCs) for new TB vaccines. We assumed the vaccines would prevent progression to disease with an average ten-years duration of protection and exponential waning. We assumed the infant vaccine would be delivered through the routine vaccination program, and the adolescent/adult vaccine delivered through routine vaccination of nine-year-olds plus a one-time vaccination campaign for ages 10+. Based on consultation with global stakeholders, we assumed a coverage target of 85% for the infant vaccine (average coverage of diphtheria-tetanus-pertussis third dose for LMICs), 80% for routine delivery of the adolescent/adult vaccine, and 70% for the adolescent/adult vaccination campaign,^14^ with equal coverage achieved within each income quintile in each country. We assumed countries would achieve linear scale-up to the specified coverage over five years and introduce vaccination in country-specific years from 2028–2047, determined based on indicators for disease burden, immunization capacity, classification of the country as an “early adopter/leader,” lack of regulatory barriers, and commercial prioritization (Appendices S1–S2).^15^

### Mathematical model

We developed a system of epidemiological and economic models, calibrated to demographic, epidemiological, and health service data in 105 LMICs (accounting for 94·4% of TB incidence in LMICs^16^)· Full epidemiological model details are described by Clark and colleagues (summarized in Appendix S1).^15^ For each country, the model was stratified by income level (lower 40% vs. upper 60% of population by household income), to reflect higher respiratory contact rates, greater TB risk factor prevalence, and poorer healthcare access among lower income groups. The risk ratio of TB disease in the low-income stratum (vs. high-income stratum) was calibrated to empirical data on income-based differentials in TB prevalence (Appendix S3).^15^

We simulated future TB-related outcomes in each modelled country for multiple scenarios over a 2028–2050 evaluation period. Each vaccine introduction scenario was compared to a ‘no-new-vaccine’ counterfactual—with current TB interventions continuing into the future at their current level—to estimate incremental changes in health service utilization and TB-related health outcomes produced by vaccine introduction.

### Health outcomes

We assessed health outcomes as incident TB cases averted. To report results by household income quintile, we further stratified the modelled income strata to create five groups of equal population size (poorest/poorer/middle/richer/richest) with TB burden in these groups following the distribution of TB cases across income strata within published TB prevalence studies (Appendix S3). For each income quintile, we aggregated results across major country groupings (global, WHO region, World Bank income level,^17^ and WHO high-TB burden grouping), to summarize the magnitude and *within-country* distribution of health gains.

### Costs incurred by TB-affected households

For each modelled country and income quintile, we calculated the costs incurred by TB patients and their households during the disease episode and applied these to the simulated number of TB cases by country, income quintile, scenario, and year. Country-specific estimates of the patient costs per TB episode were derived from a meta-regression analysis of 20 nationally-representative TB patient cost surveys.^10^ This study reported estimates for direct medical costs (medical products and services), direct non-medical costs (travel, accommodation, food, nutritional supplements), and indirect costs (income losses) incurred by TB patients, stratified by country and household income quintile, which we extracted for this analysis (Appendix S3). For each country and income quintile we assumed that the per-patient costs of TB (in 2020 constant dollars) would not change in future years. For the base-case analysis we assumed that individuals with TB disease who do not receive appropriate treatment (directly observed treatment, short-course (DOTS)), experience the same total per-episode costs as those who receive appropriate treatment, and examined alternative assumptions in sensitivity analyses. We assumed that a new TB vaccine would be provided free-of-charge and that households would incur no additional costs to receive the vaccine. We summarized results by country grouping and report costs in 2020 US dollars.

### Catastrophic costs

Following the WHO End TB target definition, we defined catastrophic costs as instances where the patient costs of TB disease—the sum of direct medical costs, direct non-medical costs, and indirect costs—exceeded 20% of total annual income for the TB-affected household.^12,18-20^ For each country and income quintile, we assessed the number of TB-affected households experiencing catastrophic costs under each scenario, multiplying the probability of catastrophic costs per TB episode by the simulated number of TB cases by country, income quintile, scenario and year. Estimates of the probability of catastrophic costs per TB episode (stratified by country and income quintile) were derived from the meta-analysis of TB patient cost surveys^10^ used for patient cost estimates (Appendix S3). For each country and income quintile, we assumed that the probability of catastrophic costs for TB patients would not change in future years. We summarized catastrophic cost results by country income-level grouping.

### Distribution of benefits across countries and income strata

We undertook additional analyses describing how each major outcome (health gains, reductions in costs faced by patients, reductions in the proportion of households experiencing catastrophic costs) was distributed across the collective income gradient of the modelled countries. First, we ordered all country income quintiles (105 countries x 5 quintiles = 525 unique groups) by average per capita gross domestic product (GDP) in 2020 purchasing power parity (PPP)-adjusted dollars. To do so we obtained estimates of per capita PPP GDP and the fraction of total income held by each country income quintile, imputing missing values according to WHO region and income level group averages (e.g., low-income countries in the African region). We multiplied these two terms and divided by the population fraction in each quintile (0.2) to obtain the average per capita PPP GDP for each quintile. We ranked all quintiles by average per-capita PPP GDP and calculated the distribution of each study outcome across these quintiles. We summarized results graphically and using the Concentration Index. The Concentration Index, defined in [-1, 1], quantifies the relative concentration of a given outcome in high- or low-income groups, with more positive (negative) values indicating greater concentration of the outcome in higher (lower) income groups (Appendix S3).

### Sensitivity analysis

We propagated uncertainty in analytic inputs using a 2^nd^-order Monte Carlo simulation (Appendix S3). This analysis generated 1000 estimates for each outcome, which we summarized as equal-tailed 95% uncertainty intervals. We also examined the robustness of results to alternative analytic assumptions. First, compared to the base-case assumption of 50% efficacy for the adolescent/adult vaccine, we examined 75% efficacy conferred by this vaccine.^15^ Second, we examined an accelerated vaccine scale-up scenario whereby all countries introduce vaccination in 2025 and achieve instantaneous scale-up to the coverage target.^15^ Third, as there is substantial uncertainty around the costs incurred by patients who do not receive TB treatment, we re-estimated results under alternative scenarios that assumed costs for this group were 50% lower and higher, respectively, compared to individuals receiving TB treatment (vs. the main analysis which assumed treated and untreated patients bore the same costs). Fourth, we examined alternative thresholds for defining catastrophic costs as 10% and 25% of total annual household income (vs. 20% in the main analysis^1^). Fifth, we examined an alternative definition of catastrophic costs that only included direct medical costs (vs. the main analysis which considered direct medical, direct non-medical, and indirect costs). Finally, we reanalysed cost results applying a 3% discount rate (vs. no discounting in the main analysis).

### Role of the funding source

The funder was involved in developing the research question, study design, and provided comments on the manuscript draft, but had no role in the collection, analysis, and interpretation of the data. All authors had the opportunity to access and verify the data, and all authors were responsible for the decision to submit the manuscript for publication.

## Results

### Overall impact of vaccine introduction

In the base-case analysis, our previous work showed that, over 2028–2050 across all 105 modelled LMICs, 6·7 (95% uncertainty interval: 5·8–7·7) million TB cases would be averted by the infant vaccine and 44·0 (37·2–51·6) million cases by the adolescent/adult vaccine, as compared to the no-new-vaccine counterfactual.^15^

In these scenarios, the infant vaccine averted costs borne by TB-affected households totalling $5907 ($5333–6533) million, including $1036 ($920–1143) million in direct medical costs, $2264 ($2027–2509) million in direct non-medical costs, and $2607 ($2351–2896) million in indirect costs (Table 1). The adolescent/adult vaccine averted $38,860 ($36,594–42,461) million in total patient costs, including $7252 ($6758–7755) million in direct medical costs, $14,987 ($13,999–16,044) million in direct non-medical costs, and $16,620 ($15,574–17,858) million in indirect costs (Table 2). We estimated 3·7 (3·3–4·1) million fewer households experiencing catastrophic costs with the infant vaccine and 22·9 (21·4–24·5) million fewer households with the adolescent/adult vaccine.

**Table 1.** Costs borne by TB-affected households averted and number of households with catastrophic costs averted by infant tuberculosis vaccines (in millions).

| Country grouping | Direct medical costs borne by TB-affected households averted | Direct non-medical costs borne by TB-affected households averted | Indirect costs borne by TB-affected households averted | Total costs borne by TB-affected households averted | Number of households with catastrophic costs averted |
| --- | --- | --- | --- | --- | --- |
| All countries | 1036 (920–1143) | 2264 (2027–2509) | 2607 (2351–2896) | 5907 (5333–6533) | 3.69 (3.31–4.08) |
| High-TB burden<sup>a</sup> | 814 (707–912) | 1992 (1763–2236) | 2112 (1876–2374) | 4917 (4368–5497) | 3.32 (2.94–3.70) |
| High-TB/HIV burden<sup>a</sup> | 668 (566–764) | 1728 (1504–1967) | 1848 (1621–2097) | 4245 (3702–4807) | 2.84 (2.50–3.21) |
| High-MDR/RR-TB burden<sup>a</sup> | 783 (677–882) | 1783 (1561–2020) | 1913 (1675–2172) | 4479 (3939–5054) | 2.89 (2.53–3.27) |
| Income level<sup>b</sup> |  |  |  |  |  |
| LIC | 181 (145–219) | 317 (270–369) | 599 (487–725) | 1096 (903–1299) | 0.61 (0.52–0.73) |
| LMIC | 699 (598–798) | 1731 (1506–1976) | 1554 (1360–1780) | 3984 (3472–4543) | 2.85 (2.50–3.25) |
| UMIC | 156 (134–179) | 217 (175–262) | 454 (334–596) | 827 (653–1030) | 0.23 (0.17–0.29) |
| World region |  |  |  |  |  |
| AFR | 483 (399–558) | 1079 (920–1229) | 1238 (1057–1425) | 2800 (2405–3182) | 1.78 (1.55–2.01) |
| AMR | 27 (23.3–30.8) | 25.8 (23–28.7) | 27.9 (24.9–31.3) | 80.8 (72.3–89.6) | 0.01 (0.01–0.02) |
| EMR | 190 (148–234) | 302 (231–379) | 513 (391–630) | 1006 (779–1239) | 0.51 (0.38–0.65) |
| EUR | 21.9 (17.6–26.6) | 10.5 (9–12.1) | 14.9 (12.3–17.5) | 47.2 (39.1–55.8) | 0.008 (0.006–0.009) |
| SEAR | 191 (144–250) | 631 (489–800) | 513 (403–655) | 1335 (1030–1710) | 1.01 (0.80–1.28) |
| WPR | 122 (103–144) | 216 (172–263) | 300 (232–374) | 639 (512–773) | 0.37 (0.28–0.47) |

Note: Values in parentheses represent equal-tailed 95% credible intervals. Total costs included patient direct medical, direct non-medical, and indirect costs (all undiscounted) in 2020 USD. Catastrophic costs are defined as instances where the patient costs (either direct medical only or total) incurred during an episode of TB disease exceeded 20% of total annual household income.
<sup>a</sup> High-TB, high-TB/HIV (HIV-associated TB), and high-MDR/RR-TB (multidrug/rifampicin-resistant TB) burden countries as defined by the World Health Organization.
<sup>b</sup> LIC: Gross national income (GNI) per capita of \$1,085 or less; LMIC: GNI per capita of \$1,086 to \$4,225; UMIC: GNI per capita of \$4,256 to \$13,205 (World Bank 2021).
Note: All countries include 105 low- and middle-income countries analysed. AFR = African region; AMR = Region of the Americas; EMR = Eastern Mediterranean region; EUR = European region; GDP = gross domestic product; LIC = low-income; LMIC = lower middle-income; SEAR = Southeast Asian region; UMIC = upper middle-income; WPR = Western Pacific region.

**Table 2.** Costs borne by TB-affected households averted and number of households with catastrophic costs averted by adolescent/adult tuberculosis vaccines (in millions).

| Country grouping | Direct medical costs borne by TB-affected households averted | Direct non-medical costs borne by TB-affected households averted | Indirect costs borne by TB-affected households averted | Total costs borne by TB-affected households averted | Number of households with catastrophic costs averted |
| --- | --- | --- | --- | --- | --- |
| All countries | 7252 (6758–7755) | 14987 (13999–16044) | 16620 (15574–17858) | 38860 (36594–41461) | 22.9 (21.4–24.5) |
| High-TB burden<sup>a</sup> | 5524 (5078–5956) | 12879 (11895–13896) | 13701 (12699–14820) | 32103 (29894–34544) | 20.2 (18.7–21.8) |
| High-TB/HIV burden<sup>a</sup> | 3807 (3449–4185) | 10719 (9803–11698) | 11507 (10547–12597) | 26033 (23851–28389) | 17.3 (15.8–18.8) |
| High-MDR/RR-TB burden<sup>a</sup> | 5614 (5163–6103) | 11676 (10733–12687) | 12588 (11605–13676) | 29878 (27590–32293) | 17.9 (16.4–19.4) |
| Income level<sup>b</sup> |  |  |  |  |  |
| LIC | 876 (770–998) | 1686 (1533–1849) | 2842 (2494–3221) | 5405 (4812–6034) | 3.31 (3.02–3.62) |
| LMIC | 4146 (3759–4547) | 10628 (9663–11593) | 9449 (8627–10315) | 24223 (22111–26293) | 17.3 (15.8–18.8) |
| UMIC | 2230 (2002–2481) | 2673 (2464–2899) | 4329 (3774–4905) | 9232 (8388–10066) | 2.25 (2.01–2.50) |
| World region |  |  |  |  |  |
| AFR | 2126 (1933–2336) | 5084 (4684–5500) | 6597 (5968–7268) | 13808 (12713–15014) | 8.52 (7.93–9.16) |
| AMR | 415 (370–461) | 530 (489–572) | 513 (472–558) | 1458 (1349–1568) | 0.30 (0.28–0.32) |
| EMR | 846 (717–988) | 1410 (1170–1651) | 2161 (1801–2557) | 4418 (3734–5169) | 2.35 (1.92–2.83) |
| EUR | 428 (364–507) | 219 (196–243) | 300 (263–341) | 947 (830–1078) | 0.15 (0.13–0.17) |
| SEAR | 1689 (1410–1997) | 5638 (4829–6510) | 4362 (3696–5071) | 11689 (9952–13516) | 8.94 (7.68–10.3) |
| WPR | 1748 (1546–1967) | 2105 (1905–2323) | 2686 (2368–3034) | 6539 (5907–7246) | 2.63 (2.29–3.04) |

Note: Values in parentheses represent equal-tailed 95% credible intervals. Total costs included patient direct medical, direct non-medical, and indirect costs (all undiscounted) in 2020 USD. Catastrophic costs are defined as instances where the patient costs (either direct medical only or total) incurred during an episode of TB disease exceeded 20% of total annual household income.
<sup>a</sup> High-TB, high-TB/HIV (HIV-associated TB), and high-MDR/RR-TB (multidrug/rifampicin-resistant TB) burden countries as defined by the World Health Organization.
<sup>b</sup> LIC: Gross national income (GNI) per capita of \$1,085 or less; LMIC: GNI per capita of \$1,086 to \$4,225; UMIC: GNI per capita of \$4,256 to \$13,205 (World Bank 2021).
Note: All countries include 105 low- and middle-income countries analysed. AFR = African region; AMR = Region of the Americas; EMR = Eastern Mediterranean region; EUR = European region; GDP = gross domestic product; LIC = low-income; LMIC = lower middle-income; SEAR = Southeast Asian region; UMIC = upper middle-income; WPR = Western Pacific region.

### Health gains by quintile

For both vaccine products, the number of TB cases averted by vaccine introduction was greatest in lower income quintiles. Across all modelled countries, the poorest two income quintiles accounted for 56% of total averted TB cases for both infant and adolescent/adult vaccination scenarios (Figure 1, Panel A), with a Concentration Index of -0·19. Figure 2 reports time trends in TB cases by income quintile for the adolescent/adult vaccination scenario, with relative differences in TB burden across income groups narrowing progressively over time (i.e., a greater relative decline in the poorest compared to the richest income quintile), in addition to the absolute reductions experienced by all groups.

**Figure 1.**
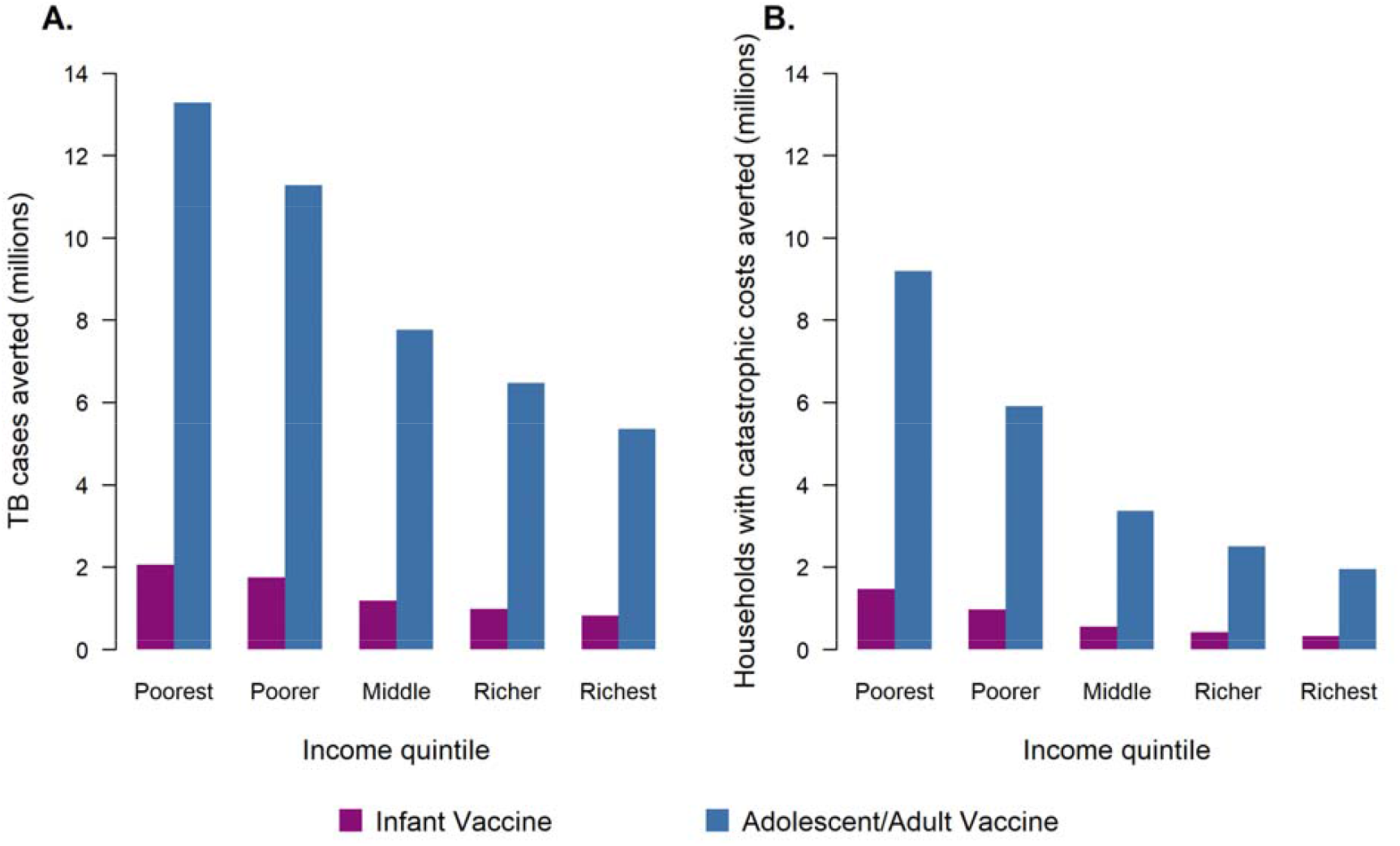
Tuberculosis cases averted (Panel A) and number of households with catastrophic costs averted (Panel B) by within-country income quintile comparing infant vaccine to adolescent/adult vaccine. Note: The cost of a TB episode presented here included patient direct medical, direct non-medical, and indirect costs. Costs borne by TB-affected households are categorized as “catastrophic” if they exceed 20% of total household’s annual income.

**Figure 2.**
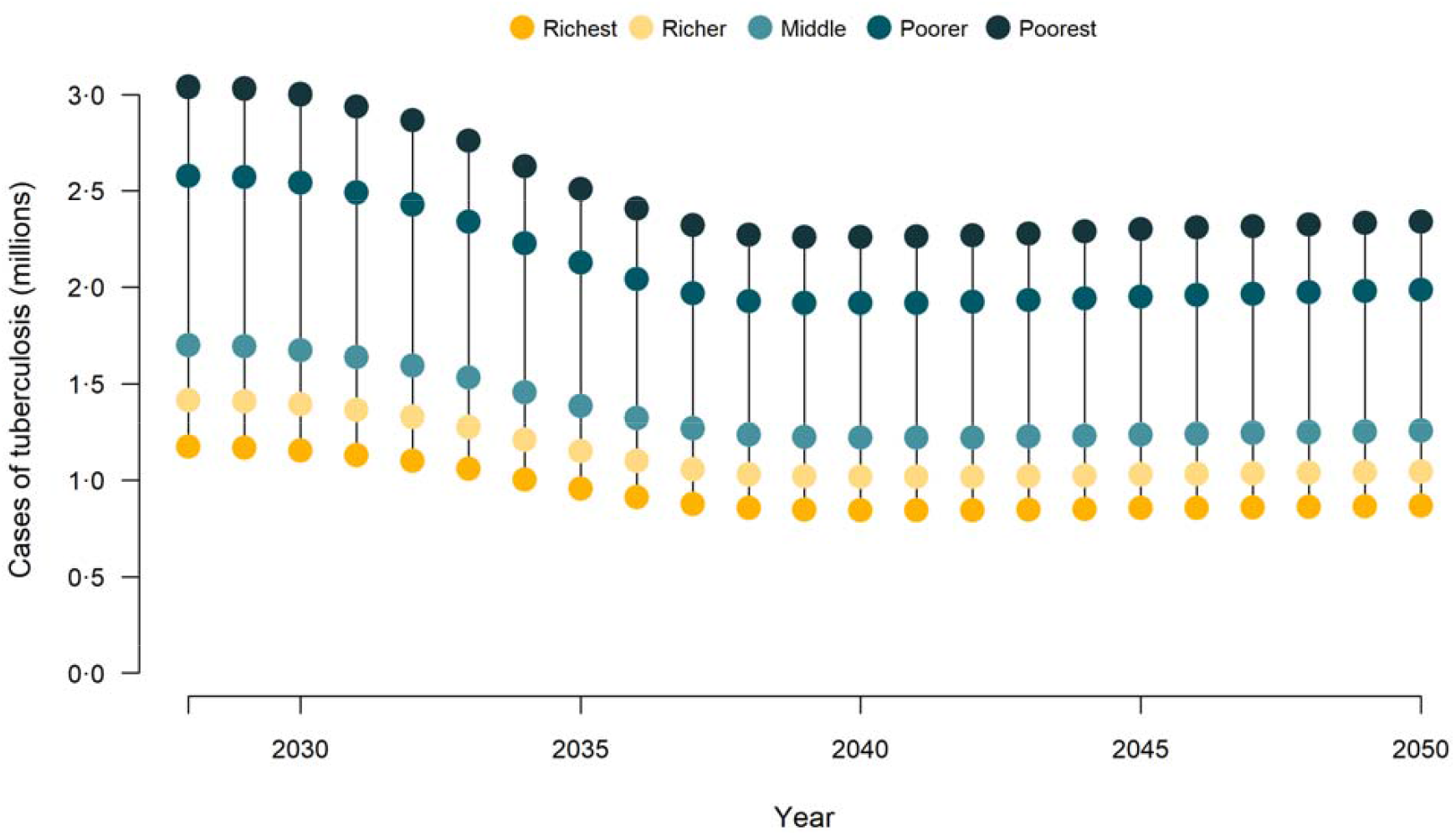
Cases of tuberculosis over time and by income quintile with delivery of adolescent/adult tuberculosis vaccines across 105 low- and middle-income countries. Note: Country-specific vaccine introduction years from 2028–2047.

### Averted patient costs by income quintile

The absolute reductions in TB patient costs resulting from vaccine introduction were weighted slightly towards higher income quintiles, with greater costs per episode of TB care incurred in these groups (Appendix S4) outweighing the greater reduction in TB cases in poorer quintiles. Across all modelled countries, the wealthiest two income quintiles accounted for 45% of total averted patient costs for the infant vaccination scenario (Concentration Index 0·06), and 46% for the adolescent/adult vaccination scenario (Concentration Index 0·07). When results were disaggregated by cost category, direct medical costs averted showed relatively equal distribution across quintiles (Concentration Index 0·03 and 0·04 for infant and adolescent/adult vaccines, respectively), while direct non-medical costs averted were concentrated in the poorest two quintiles (Concentration Index -0·09 and -0·08 for infant and adolescent/adult, respectively). The majority (56%) of indirect costs averted were concentrated in the wealthiest two quintiles (Concentration Index 0·20 and 0·21 for infant and adolescent/adult, respectively).

### Catastrophic costs averted by income quintile

The largest absolute reductions in the proportion of households facing catastrophic costs were in lower income quintiles within each country. Under each vaccination scenario, 66% of cases of catastrophic costs were averted in the poorest two quintiles (Concentration Index -0·31). Figure 1, Panel B shows the relative magnitude of cases of catastrophic costs averted across income quintiles. This gradient is steeper than for TB cases averted (Panel A), indicating greater differences by quintile for catastrophic costs.

### Distribution of benefits across countries and income strata

Figure 3 shows the distribution of TB cases and catastrophic costs averted by household income across the combined population of the modelled countries over the 2028–2050 period for the adolescent/adult vaccine compared to the no-new-vaccine counterfactual. For both outcomes the benefits of vaccine introduction were concentrated in poorer households (poorest quintile shaded in red), with 18·3 (14·2–22·7) million TB cases projected to be averted in the poorest 20% of households (41% of total cases averted, Concentration Index -0·36), and 12·1 (9·4–15·0) million cases of catastrophic cost averted in the poorest 20% of households (53% of total cases of catastrophic costs averted, Concentration Index -0·48). Reductions in patient costs were also greater in the poorest households (Appendix S5), with $11·8 (9·1–14·6) billion in cost savings projected to be averted in the poorest 20% of households (30% of total patient costs averted, Concentration Index -0·15).

**Figure 3.**
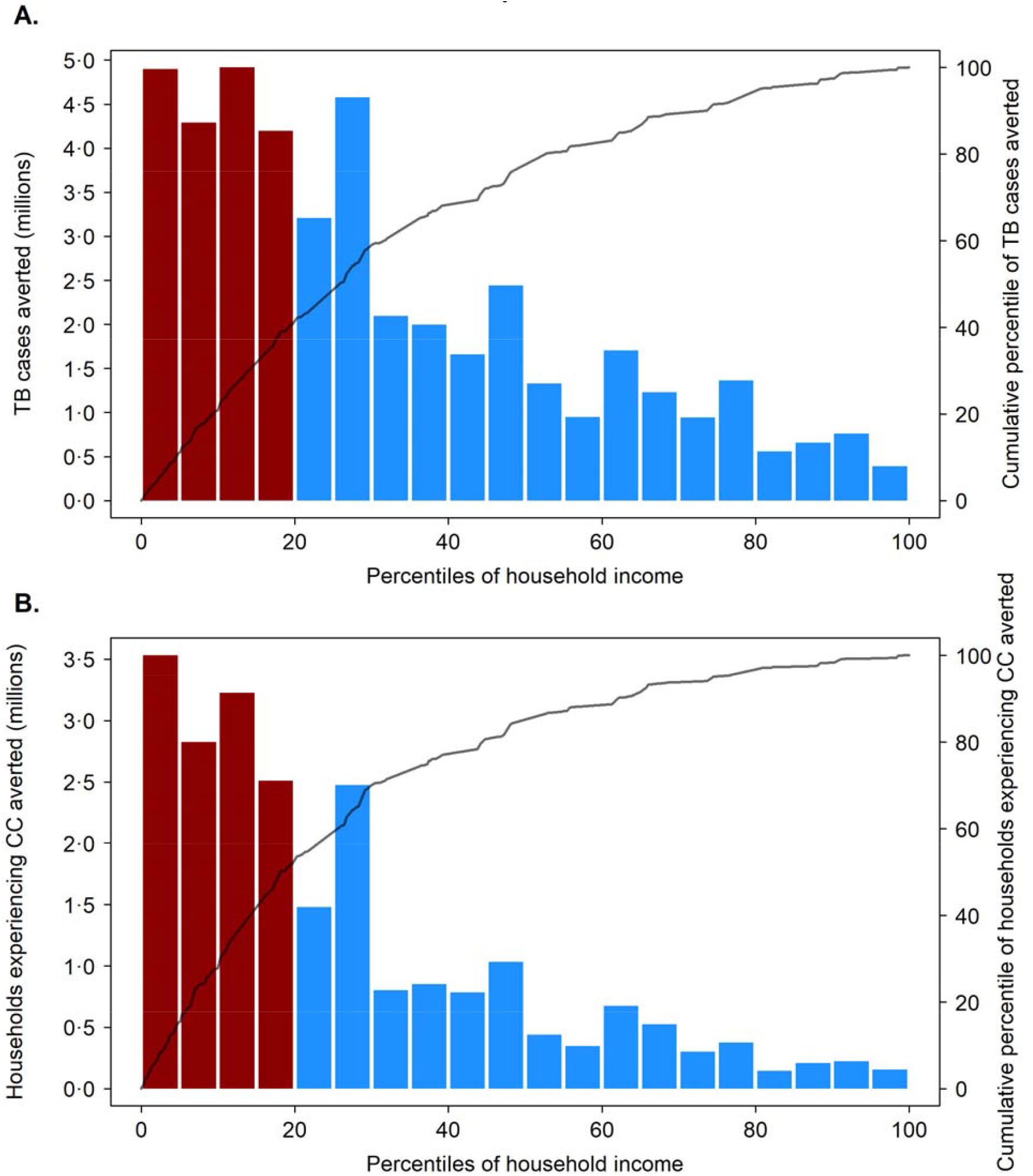
Distribution of tuberculosis cases averted (Panel A) and number of households experiencing catastrophic costs averted over 2028–2050 (Panel B) by an adolescent/adult vaccine across all modelled strata, ordered by household income. Note: TB = tuberculosis, CC = catastrophic costs; GDP = gross domestic product. Bars defined by left-hand side y-axis; lines defined by right-hand side y-axis. Ordering of population by household income based on average 2020 per-capita GDP in purchasing power parity (PPP) dollars, for each modelled stratum (505 total strata). Bars shaded red indicate poorest 20% of modelled population by PPP GDP per capita.

### Sensitivity analyses

Compared to the assumed base-case efficacy of 50% for the adolescent/adult vaccine, an assumption of 75% efficacy had greater impact, averting 33·4 (31·2–35·8) million cases of catastrophic costs (Appendix S6). Compared to the base-case scenario, the accelerated scale-up scenario had greater household economic impact for both infant and adolescent/adult vaccination (Appendices S7–S8), with 8·8 (8·0–9·8) million cases of catastrophic costs averted for the infant vaccine and 33·9 (31·7–36·3) million for the adolescent/adult vaccine. Assuming that individuals with TB who do not receive appropriate treatment experience 50% lower costs compared to treated individuals reduced the number of households with averted catastrophic costs, to 2·8 (2·5–3·1) million for the infant vaccine and 18·7 (17·6–20·0) million for the adolescent/adult vaccine (Appendices S9–S11). Assuming these individuals experience 50% higher costs compared to treated individuals increased the number of households with averted catastrophic costs, to 4·2 (3·8–4·7) million for the infant vaccine and 25·4 (23·6–27·2) million for the adolescent/adult vaccine (Appendices S12–S16). Using alternative definitions of catastrophic costs, such as instances where patient costs exceed 10% of annual household income (vs. 20% in the main analysis), resulted in approximately 40% more cases of catastrophic costs averted in infant and adolescent/adult vaccine scenarios (Appendix S17). A higher threshold for defining catastrophic costs (i.e., >25% of annual household income), resulted in approximately 14% fewer cases of catastrophic costs averted in infant and adolescent/adult vaccine scenarios (Appendix S18). When we restricted the definition of catastrophic costs to only consider direct medical costs (vs. the sum of direct medical, direct non-medical, and indirect costs in the main analysis), we estimated approximately 84% fewer cases of catastrophic costs averted compared to the main analysis, for both vaccine products (Appendices S17–S18). When we applied a 3% discount rate to total (direct and indirect) costs incurred in future years (vs. no discount rate in the main analysis), total patient costs averted were $3584 ($3241–3953) million and $26,352 ($24,817– 28,081) million for infant and adolescent/adult vaccines respectively, 39% and 32% less than in the main analysis (Appendices S19–S20).

## Discussion

In this study we estimated the potential impact of new TB vaccines on income-based disparities in the health and economic consequences of TB in LMICs. Both infant and adolescent/adult vaccines were projected to reduce disparities in TB disease burden, with the adolescent/adult vaccine estimated to have greater absolute impact. Across all modelled countries over 2028– 2050, an adolescent/adult vaccine was projected to reduce the number of incident TB cases in the poorest quintile by 13·3 (10·9–15·8) million (30% of total TB cases averted) and reduce the number of households experiencing total costs of care exceeding 20% of household income (i.e., catastrophic costs) by 9·2 (7·5–11·0) million in the poorest quintile (40% of total cases of catastrophic costs averted) compared to the no-new-vaccination baseline.

The concentration of vaccine impact in lower income groups results from two features of TB in LMICs. Firstly, current TB burden is concentrated in low-income groups. Individuals in poor households are more likely to be infected, have a greater concentration of risk factors for developing TB disease, and are more likely to die from TB if it occurs.^1^ These differences are rooted in the socioeconomic determinants of TB and have motivated a global TB strategy that emphasises vulnerable groups and strengthening social protection.^21,22^ Developing an effective TB vaccine is a key component of this strategy. Secondly, poor households are uniquely vulnerable to health shocks, which can lead to economic hardship and medical debt. In the meta-analysis of TB patient cost surveys^10^ used for this analysis, the fraction of patients experiencing catastrophic costs was higher for each successive income quintile from richest to poorest.

In addition to reporting the distribution of heath and economic benefits of TB vaccination within each country, we also analysed the distribution of benefits across the combined income gradient of the 105 modelled countries. The results from this analysis are qualitatively similar to the within-country analyses,^10^ though with greater concentration of benefits within poorer groups for each outcome (more negative Concentration Index values), highlighting the concentration of potential benefits within poor countries, as well as within poor groups within each country.

This analysis has several limitations. Firstly, the characteristics of a new TB vaccine, once available, may differ from the scenarios we examined, which were based on the WHO PPCs.^13^ These PPCs represent preferred vaccine attributes to deliver public health impact, but a final product may differ in terms of effectiveness or duration of protection.^15^ Secondly, we made assumptions about vaccine introduction based on expert opinion and historical vaccine introduction patterns. In practice, the impact of vaccination will depend on how aggressively countries scale-up a new vaccine, with slow or delayed introduction reducing the magnitude of impact.^15^ Thirdly, we assumed that vaccine coverage would be the same across all income quintiles in each country. However, evidence for routine immunization delivery shows heterogeneity in coverage between income groups, with a trend towards lower coverage in poorer income quintiles.^23^ Efforts to lower access barriers in low-income groups may be needed to prevent inequalities in TB vaccine coverage. Related to this, we assumed vaccines would be provided free-of-charge. Requiring payment for vaccination would likely reduce uptake, particularly within low-income groups. Fourthly, while cost data are available for patients treated for TB, no information was available for individuals not receiving treatment. We assumed that these untreated individuals experienced the same costs as treated individuals in the base-case analysis and examined this assumption in sensitivity analysis. Finally, we assumed that TB trends in the no-new-vaccine base-case would follow their historical trajectory. If there were aggressive scale-up of non-vaccine interventions (as envisaged by recent global TB strategy), this would reduce the incremental impact of a new vaccine.^15^

## Summary

Policymakers consider several issues when prioritizing health interventions, one of which is impact on health equity.^24^ This manuscript demonstrates that under a range of assumptions, new TB vaccines could be highly impactful and help narrow income-based disparities in the health and the economic consequences of TB for low- and middle-income countries, helping achieve the WHO End TB Strategy goals, make substantial progress towards achieving Universal Health Coverage, and sustainable development goal targets (e.g., eradicating poverty (SDG 1), hunger (SDG 2), promoting decent work and growth (SDG 8), and good health and well-being (SDG 3). To achieve these benefits, countries will need to commit to rapid introduction once an effective vaccine is approved, achieve high population coverage, and prevent differentials in vaccine access for poor and marginalized groups. Doing so will require sustained political and financial commitments by affected countries and international partners, as well as implementation research on approaches to eliminate access barriers during vaccine introduction. While major challenges remain, successful development and introduction of a new TB vaccine has the potential to accelerate the elimination of a disease that has represented one of the greatest health threats for poor households for millennia.

## Data Availability

All data produced in the present study are available upon reasonable request to the authors.

## Acknowledgements

We are grateful for the support of the World Health Organization for funding this research (2020/985800-0). We thank all the attendees at the WHO meetings on the Full Value Assessment of TB Vaccines for insightful advice and direction.

## Ethics approval and consent to participate

Not applicable.

## Author contributions

AP, NAM, RAC, MQ, MJ, and RGW conceptualized and designed the study. AP conducted the analysis with support from NAM, RAC, MQ, MJ, CKW, CM, and RGW. AP and NAM drafted the initial manuscript and approved the final manuscript as submitted. MQ, CKW, CM, RB, NG, MZ, IGB, NN, and MJ critically reviewed the analysis, reviewed and revised the manuscript, and approved the final manuscript as submitted. All authors read and approved the final manuscript.

## Declarations of interests

RAC is funded by BMGF (INV-001754) and received a grant from the Canadian Centennial Scholarship Fund. CKW is funded by UKRI/MRC (MR/N013638/1). RGW is funded by the Wellcome Trust (218261/Z/19/Z), NIH (1R01AI147321-01), EDTCP (RIA208D-2505B), UK MRC (CCF17-7779 via SET Bloomsbury), ESRC (ES/P008011/1), BMGF (OPP1084276, OPP1135288 & INV-001754), and the WHO (2020/985800-0). All other authors declare no conflicts of interest.

## Notes

### Funding Statement

This study was funded by World Health Organization (2020/985800-0).

