## Supplementary Appendix for "The potential impact of novel tuberculosis vaccines on health equity and financial protection in low- and middle-income countries"

**Appendix S1. Epidemiological methods**

The subsequent pages provide relevant details and methods regarding the underlying epidemiological model and vaccine delivery scenarios from Clark et al.^1^

**S1.1.	Tuberculosis natural history structure**

The core natural history model is specified in Figure S1. Those with no previous exposure or infection with *Mtb* [Uninfected-Naive (U_N_)] could become infected at rate
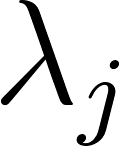
 and progress to an Infection-Fast (I_F_) class following initial infection. From Infection-Fast, three possible pathways were possible: (i) Fast progression to Subclinical Disease (D_S_), where individuals are infectious with a reduced infectiousness compared to clinical tuberculosis, but display no symptoms of tuberculosis disease;^2^ (ii) self-clearance to Uninfected-Cleared (U_C_), where individuals are no longer infected with *Mtb* and therefore are not at risk of progression to tuberculosis disease without reinfection;^3^ or (iii) continue to remain latently infected with a risk of reactivation and progression to disease, albeit at a lower rate than Infection-Fast, by transitioning to the Infection-Slow (I_S_) class. Those in the Infection-Slow class could self-clear to the Uninfected-Cleared class, be reinfected and return to the Infection-Fast class or reactivate their infection and progress to Subclinical Disease.

Once in the Subclinical Disease class, individuals could naturally cure (without treatment) to the Resolved (R) class, or progress to Clinical Disease (D_C_), where individuals are infectious and display symptoms of tuberculosis disease. Treatment initiation from Clinical Disease to On-Treatment (T) began in 1960 and increased following a sigmoid curve to 2019, with average treatment duration assumed to be six months.^4,5^ Treatment completions transitioned to the Resolved class and treatment non-completions returned to Clinical Disease. Deaths occurring on-treatment and in clinical disease counted toward the total number of tuberculosis deaths during the year. Those with clinical disease could also naturally cure to the resolved class. Individuals in the Resolved class could be reinfected or relapse to Subclinical. We assumed that the infection and resolved classes are partially protected against reinfection.^6,7^ In those who have self-cleared, we assumed the level of protection against reinfection is half of the protection against reinfection for the infection and resolved classes. Age was modelled in single years from ages 0 to 79 and aggregated into two categories for ages 80 to 89, and ages 90 to 99. Births and ageing occurred at the beginning of each year.

**S1.2. HIV and ART structure description**

To account for the influences of human immunodeficiency virus (HIV) and antiretroviral therapy (ART) on the risk of infection with *Mtb* and progression to tuberculosis disease,^6,8^ we implemented an HIV structure (shown in Figure S2) composed of 3 compartments: HIV uninfected [HIV0], people living with HIV (PLHIV) not on ART [HIV1], and PLHIV on ART [ART]. HIV uninfected individuals were diagnosed with HIV and moved from the HIV0 compartment to the HIV1 compartment with rate
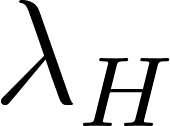
<https://www.codecogs.com/eqnedit.php?latex=%5Clambda_H#0>. Within the HIV1 compartment, there is a higher risk of tuberculosis progression and an increased tuberculosis mortality rate compared to the HIV0 compartment. PLHIV are initiated on treatment with ART from HIV1 following a sigmoid trend. The increases in tuberculosis mortality rate and tuberculosis progression are reduced while in ART compared to HIV1, but still higher than in HIV0. ART also reduces the HIV mortality rate.

The separate stratum was included to dynamically model the tuberculosis-HIV co-epidemic if the proportion of tuberculosis cases among people living with HIV (PLHIV) was greater than or equal to 15%, and if the HIV prevalence in the country was greater than 1%. Countries incorporating the additional HIV structure are listed in Table S3.

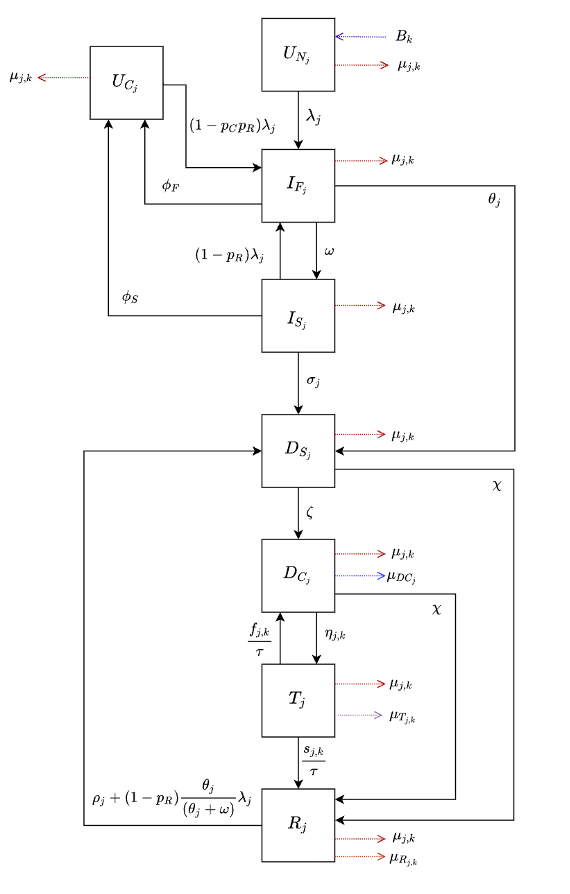

**Figure S1.	Tuberculosis natural history model**

*Abbreviations: D_C_ = Clinical Disease; D_S_ = Subclinical Disease; I_F_ = Infection-Fast; I_S_ = Infection-Slow; R = Resolved; T = On-Treatment; U_C_ = Uninfected-Cleared; U_N_ = Uninfected-Naïve.*

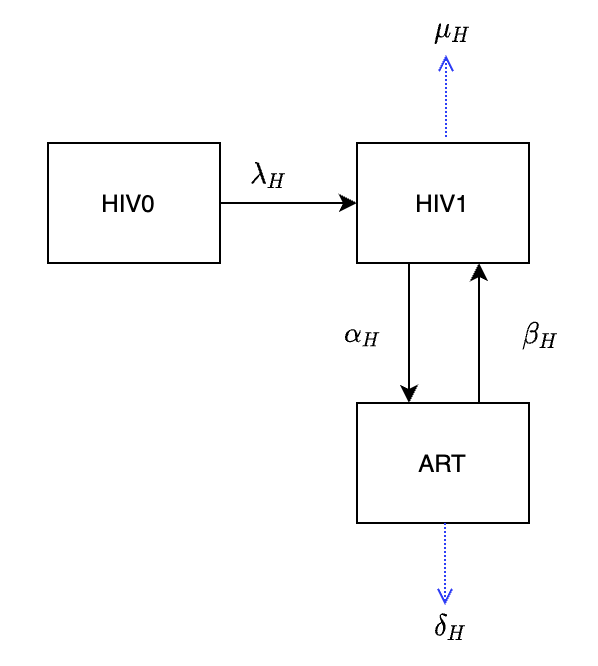

**Figure S2.	HIV and ART structure.**

*Abbreviations: ART = People living with HIV on ART; HIV0 = HIV uninfected; HIV1= People living with HIV not on ART.*

**Table S3.**	**Countries incorporating the HIV structure with their corresponding HIV prevalence and proportion of tuberculosis cases among PLHIV.**

| **Country** | **HIV Prevalence (%)** | **Proportion of tuberculosis cases among PLHIV (%)** |
| --- | --- | --- |
| Botswana | 16·5 | 48·6 |
| Central African Republic | 2·1 | 25·4 |
| Côte d’Ivoire | 1·7 | 17·5 |
| Cameroon | 2·0 | 26·8 |
| Gabon | 2·3 | 32·8 |
| Ghana | 1·1 | 20·8 |
| The Gambia | 1·2 | 17·7 |
| Guinea-Bissau | 2·1 | 31·3 |
| Equatorial Guinea | 4·8 | 26·5 |
| Guyana | 1·1 | 19·0 |
| Kenya | 2·9 | 26·2 |
| Lesotho | 16·0 | 61·6 |
| Mozambique | 7·2 | 33·8 |
| Malawi | 5·9 | 46·6 |
| Namibia | 8·4 | 32·5 |
| Rwanda | 1·8 | 21·1 |
| Eswatini | 17·4 | 60·1 |
| Togo | 1·5 | 16·2 |
| Tanzania | 2·9 | 23·6 |
| Uganda | 3·4 | 39·0 |
| South Africa | 12·8 | 58·0 |
| Zambia | 6·7 | 46·2 |
| Zimbabwe | 9·6 | 59·8 |

**S1.3. Access to care structure**

The access to care dimension is incorporated to allow for the negative correlation between tuberculosis burden and health care access to prevent the overestimation of vaccine impact, as well as to facilitate future analyses of equity implications of vaccine introduction. The access to care dimension contains 2 classes: high-access-to-care, representing the top 3 quintiles (60% of the population) and low-access-to-care, representing the bottom 2 quintiles (40% of the population). We assumed that there was no transition between the high- and low-access-to-care classes, as well as assuming random mixing between the high-access-to-care and low-access-to-care classes.

To constrain relative burden between access-to-care classes, we calibrated the relative tuberculosis prevalence in the high-access-to-care class to the low-access-to-care class in 2019. The calibration target, 0·674, was calculated as a weighted average from eleven studies, with lower and upper bounds (0·575–0·801) representing the 25th and 75th percentiles of the datasets (Table S4).^9-19^

**Table S4.** TB prevalence study data.

| Source | Country | Prevalence rate ratio of upper 60% vs. lower 40% of population by socioeconomic status | Weight |
| --- | --- | --- | --- |
| ^11^ | Bangladesh | 0·394 | 1 |
| ^14^ | India | 0·386 | 1 |
| ^15^ | India | 0·467 | 1 |
| ^17^ | Kenya | 0·588 | 1 |
| ^16^ | Malawi | 0·867 | 1 |
| ^16^ | Mongolia | 0·716 | 1 |
| ^16^ | Myanmar | 0·807 | 1 |
| ^16^ | Philippines | 0·755 | 0·5 |
| ^17^ | Philippines | 0·608 | 0·5 |
| ^13^ | Rwanda | 1·081 | 1 |
| ^16^ | Rwanda | 0·774 | 1 |
| ^9^ | South Africa | 0·486 | 1 |
| ^18^ | South Africa | 0·896 | 1 |
| ^16^ | Tanzania | 0·648 | 1 |
| ^10^ | Vietnam | 0·701 | 1 |
| ^19^ | Vietnam | 0·799 | 1 |
| ^16^ | Vietnam | 0·672 | 1 |
| ^12^ | Zambia | 0·534 | 0·7 |
| ^16^ | Zambia | 1·312 | 0·7 |
| ^18^ | Zambia | 0·728 | 0·7 |

To incorporate access to care into our model, we assume that the differences in tuberculosis burden between classes are due to differences in the force of infection, the rate of care-seeking (i.e., tuberculosis treatment initiation), and the rate of tuberculosis progression. We assume relative to the low-access-to-care stratum, the high-access-to-care stratum has a reduced force of infection per contact, an increased rate of treatment initiation, and a reduced rate of tuberculosis progression. Differential burden was implemented by introducing a new parameter
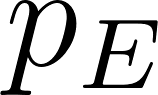
<https://www.codecogs.com/eqnedit.php?latex=p_E#0>, such that
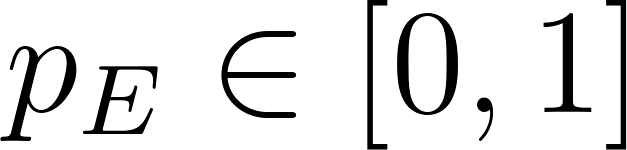
<https://www.codecogs.com/eqnedit.php?latex=p_E%20%5Cin%20%5B0%2C1%5D#0> for the high-access-to-care and
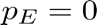
<https://www.codecogs.com/eqnedit.php?latex=p_E%20%3D%200%20#0> and included within the model natural history structure as described in Table S5. This new parameter was fitted during calibration.

**Table S5.**	Implementing the access-to-care parameter

|  | **Access-to-Care** |
| --- | --- |
| **Force of infection** | 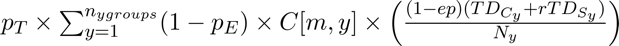<https://www.codecogs.com/eqnedit.php?latex=p_T%20%5Ctimes%20%5Csum_%7By%3D1%7D%5E%7Bn_%7Bygroups%7D%7D(1-p_E)%20%5Ctimes%20C%5Bm%2Cy%5D%20%5Ctimes%20%5Cleft%20(%20%5Cfrac%7B(1-ep)(TD_%7BC_y%7D%2BrTD_%7BS_y%7D)%7D%7BN_y%7D%20%5Cright%20)#0> |
| **Treatment Initiation Rate** | 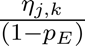<https://www.codecogs.com/eqnedit.php?latex=%5Cfrac%7B%5Ceta_%7Bj%2Ck%7D%7D%7B(1-p_E)%7D#0> |
| **Rate of Tuberculosis Progression** | 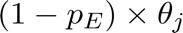<https://www.codecogs.com/eqnedit.php?latex=(1-p_E)%20%5Ctimes%20%5Ctheta_j#0>  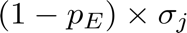<https://www.codecogs.com/eqnedit.php?latex=(1-p_E)%20%5Ctimes%20%5Csigma_j#0>  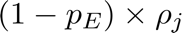<https://www.codecogs.com/eqnedit.php?latex=(1-p_E)%20%5Ctimes%20%5Crho_j#0> |

**S1.4.	Calibration methodology**

The model was fitted to epidemiologic calibration targets using history matching with emulation, implemented using the hmer R package.^20,21^ If countries were unable to find at least 1000 fully fitted parameter sets using this method, they were subsequently assessed using an Approximate Bayesian Computation using Markov Chain Monte Carlo method (ABC-MCMC). ABC-MCMC was conducted using the easyABC package in R, modified by Sebastian Funk, Gwenan Knight, and the Tuberculosis Modelling group at LSHTM for adaptive sampling and to accept seeded parameter values.^20,22^ We used parameter sets with the maximum number of targets fitted using history matching with emulation as a starting seed, with the ABC-MCMC algorithm continuously adapting using the last 1000 points and the noise factor set to 0·0001.

Analysis was performed on 105 countries from the 135 total low- and middle-income countries identified based on 2019 World Bank Income groups. There were 20 countries excluded from the initial calibration attempt due to missing crucial data required to attempt calibration, and 10 countries which were unable to be calibrated (could not find a parameter set that matched all targets using both history matching with emulation as well as ABC-MCMC). Reasons for exclusion from the final list of calibrated countries are provided in Table S6.

**Table S6.	Reasons for exclusion from the final list of calibrated countries.**

| **Country** | **Reason for Exclusion** |
| --- | --- |
| Algeria | Did not calibrate |
| American Samoa | Missing multiple critical epidemiological data for calibration, no contact matrices available |
| Belize | No case notification or incidence data for children |
| Bosnia and Herzegovina | Did not calibrate |
| Cabo Verde | Did not calibrate |
| Comoros | No case notification data |
| Democratic Republic of the Congo | No case notification data by age |
| Republic of the Congo | No population estimates |
| Democratic People's Republic of Korea | Missing multiple critical epidemiological data for calibration |
| Djibouti | No case notification data by age |
| Dominica | Missing multiple critical epidemiological data for calibration, no contact matrices available |
| Guinea-Bissau | Did not calibrate |
| Guyana | Did not calibrate |
| Federated States of Micronesia | Missing multiple critical epidemiological data for calibration, no contact matrices available |
| Grenada | Missing multiple critical epidemiological data for calibration, no contact matrices available |
| Haiti | Missing 2020 contact matrix |
| Jamaica | Did not calibrate |
| Kiribati | Missing 2020 contact matrix |
| Kosovo | Missing multiple critical epidemiological data for calibration |
| Lebanon | Missing 2020 contact matrix |
| Marshall Islands | Missing multiple critical epidemiological data for calibration, no contact matrices available |
| North Macedonia | Did not calibrate |
| Samoa | No case notification or incidence data for children |
| Somalia | No contact matrices available |
| St. Lucia | No case notification or incidence data for children |
| St. Vincent and the Grenadines | Did not calibrate |
| Tonga | Did not calibrate |
| Turkmenistan | Did not calibrate |
| Tuvalu | No contact matrices available |
| West Bank and Gaza | Missing multiple critical epidemiological data for calibration |

**S1.5. Vaccine profile**

The vaccine profile for an adult/adolescent vaccine and infant vaccine were based on the WHO Preferred Product Characteristics for New Tuberculosis vaccines,^23^ and are outlined in Table S7 below.

**Table S7.**	**WHO Preferred Product Characteristics for New Tuberculosis Vaccines.**

| **Vaccine** | **Host infection status at time of vaccination required for efficacy** | **Effect type** | **Vaccine efficacy** | **Duration of protection** |
| --- | --- | --- | --- | --- |
| Adolescent / Adult | Pre- and post-infection | Prevention of disease | 50% | Lifelong |
|  |  |  |  | 10 years |
| Infant | Pre-infection | Prevention of disease | 80% | Lifelong |
|  |  |  |  | 10 years |

Vaccine efficacy was assumed to be the same in both PLHIV and HIV-naïve recipients in countries incorporating the HIV structure, and in both younger age groups and older adults. The vaccine was assumed to have the same impact on preventing drug-susceptible and drug-resistant tuberculosis as specified in the WHO PPCs.^23^ As we were modelling a prevention of disease vaccine, there was no direct impact on *Mtb* transmission or the force of infection.

We assumed duration of protection was 10 years on average, in addition to a sensitivity analysis with lifelong duration of protection. The shape of waning immunity was modelled as an exponential distribution, based on similar shapes for waning vaccine immunity of BCG^24^ and other vaccines.^25,26^

**S1.6.	Vaccine delivery scenarios**

The infant vaccine was implemented in two scenarios, and, separately, the adolescent/adult vaccine was implemented in three scenarios. The *Basecase* and *Accelerated Scale-up* scenarios included routine single-dose neonatal vaccination for the infant vaccine (85% coverage), and routine single-dose vaccination of 9-year-olds (80% coverage) with a one-time vaccination campaign for ages ten and older (70% coverage) for the adolescent/adult vaccine. The *Routine Only* scenario (adolescent/adult vaccine only) was introduced through routine 9-year-old vaccination only (i.e., no campaign). Specifics of the infant and adolescent/adult vaccine scenarios are provided in Table S8.

**Table S8.	Vaccine scenarios for the infant and adolescent/adult vaccines**.

| **Characteristics** | **Infant Vaccine Scenarios** | | **Adolescent/Adult Vaccine Scenarios** | | |
| --- | --- | --- | --- | --- | --- |
|  | ***Basecase*** | ***Accelerated***  ***Scale-up*** | ***Basecase*** | ***Accelerated***  ***Scale-up*** | ***Routine Only*** |
| **Ages Targeted** | *Neonatal:*  Routine | *Neonatal:*  Routine | *Age 9:* Routine  *Ages 10+:* One-time vaccination campaign over 5 years | *Age 9:* Routine    *Ages 10+:* One-time vaccination campaign in 2025 | *Age 9:* Routine |
| **Introduction Year** | Country-specific | 2025 | Country-specific | 2025 | Country-specific |
| **Vaccine Rollout  Trend** | 5-year linear scale-up to coverage | Instant scale-up to coverage | 5-year linear scale-up to coverage | Instant scale-up  to coverage | 5-year linear scale-up to coverage |
| **Target Coverage *(Low/Med/High)*** | 75% / 85% / 95% | | Age 9: 70% / 80% / 90%  Ages 10+: 50% / 70% / 90% | | |

**S1.7. Country-specific introduction years**

In the *Basecase* and *Routine Only* scenarios, vaccines were introduced in country-specific introduction years between 2028 and 2047. The year 2028 was selected as the earliest country-specific introduction year to align with the anticipated completion and availability of results from TB vaccine candidate trials based on expert consultation and analysis. Country-specific introduction years were calculated for all 135 LMICs based on the 2019 World Bank Income groups. To calculate the specific year of introduction, countries were divided into two general categories: those procuring with support from Gavi, the Vaccine Alliance, and those self-procuring. Determination of country status was based on eligibility information posted on Gavi’s website.^27^ Countries transitioning from Gavi support are able to benefit from Gavi pricing and incremental financing for a period of 5–10 years. For countries that have already initiated the period of transition by 2019, this window will have largely ended by the time of tuberculosis vaccine availability through Gavi. As such, these countries were categorised as self-procuring countries. Countries that have not yet commenced transition, including India and Nigeria, were categorised as Gavi supported countries, given the long grace period post-commencement of transition. For more information, please see Gavi, <https://www.gavi.org/types-support/sustainability/transition> (retrieved December 1, 2020).

Through a consultative process with experts from WHO, Gavi, PATH, PDVAC, CHAI, and industry partners, factors influencing likelihood of being an early or late adopter were identified for both Gavi and self-procuring countries. Identified factors include disease burden, immunization capacity, and early adopter status. Country-specific registration timelines and commercial prioritization were also deemed important determinants of introduction timing for self-procuring countries.

*Additional factors for Gavi countries:* For countries procuring through Gavi, timelines for introduction are also influenced by Gavi processes. Prior to offering a new vaccine, Gavi requires that products be licensed, included in Gavi’s Vaccine Investment Strategy, reviewed by SAGE, recommended in a WHO position paper, WHO prequalified, and approved for procurement by Gavi (Table S9). In addition, time for country application processing, contracting, and delivery must be factored. Through consultations, it was determined that a baseline time of roughly two years post licensure would be needed for Gavi processes prior to first country introduction, assuming several steps advance in parallel.

**Table S9.	Timelines for Gavi processes post licensure.**

|  | **Cumulative additional time (years)** | | |
| --- | --- | --- | --- |
| **Activities post licensure** | **Low End** | **High End** | **Average** |
| WHO PQ | 0·25 | 1·00 | 0·63 |
| SAGE Policy Review & WHO Position Paper | 0·25 | 0·50 | 0·38 |
| Gavi Decision | 0·25 | 0·50 | 0·38 |
| National review & Country applications | 0·25 | 0·75 | 0·50 |
| Contracting & delivery | 0·25 | 0·50 | 0·38 |
| **Years** | **1·25** | **3·25** | **2·25** |

*Weight of criteria, indicators, and scoring*: Differential weight was assigned to criteria based on their relative impact on the order of country adoption. This weight varied for self-procuring and Gavi countries (Table S10).

**Table S10.	Weight of criteria influencing order of country adoption.**

| **Criteria** | **Self-procuring countries** | **Gavi countries** |
| --- | --- | --- |
| Disease burden | 30% | 45% |
| Immunization capacity | 15% | 30% |
| Early adopter/leader | 15% | 25% |
| Lack of regulatory barriers | 15% | NA |
| Commercial prioritization | 25% | NA |

The following indicators were used to measure each of the variables identified in Table S11.

**Table S11.	Indicators of criteria influencing order of country adoption.**

| **Criteria** | **Indicator** |
| --- | --- |
| **Disease burden** | Tuberculosis incidence |
| **Immunization capacity** | Proportion receiving 3 doses of DPT3 among infants 1 years of age (The percent of infants receiving 3 doses DPT3 is commonly used as a proxy for assessing immunization infrastructure) |
| **Lack of regulatory barriers** | Signatories to WHO PQ or SRA collaborative registration scheme  Lack of requirements for additional local clinical trial data |
| **Early adopter/leader** | Time to policy adoption of universal Xpert MTB/RIF screening for presumed tuberculosis cases  Time to adoption of HPV |
| **Commercial prioritization** |  |
| *Ability to finance vaccines* | GDP per capita |
| *Political will to address tuberculosis* | Spending per tuberculosis case |
| *Market potential* | Population |

To standardize across these varied metrics, a point value ranging from 1–5 per criteria was assigned, with a score of 1 correlating with an earlier adopter and score of 5 correlating with a later adopter.

*Continuous variable*s such as disease burden or population were divided into quintiles. Those in the highest quintile were assigned a score of 1, those in the second highest quintile received a score of 2, and so forth. *Categorical variables* such as registration or early adopter status were scored based on whether countries met fixed criteria. For instance, countries that are signatories of WHO PQ or SRA collaborative registration schemes were assigned a score of 1. Those that are not signatories and have requirements for additional clinical trial data in local populations received a score of 5.

Scores were then weighted as reflected in Table S10 and aggregated into a composite score to determine countries’ relative position in the queue of introductions.

Assumptions for the pace of introduction—i.e., how many countries per year would introduce the product and what the scale up curve might look like— was informed with data from pneumococcal vaccine (PCV) scale-up.^28^ The percent of countries adopting each year (year 1 to year 12) for PCV was calculated. These annual percentages were then applied to tuberculosis vaccine scale up (based on a total n=135 countries: 78 self-procuring countries and 57 Gavi countries). The first year of tuberculosis vaccine scale up was estimated to be 2028, with Gavi countries following a similar scale up trajectory but delayed by two years due to required Gavi lead time for processing new vaccines (Table S9). Because PCV data is only available for 12 years, data was extrapolated for years 13 to 20 of tuberculosis vaccine roll out at a steady state. Country introduction timelines were adjusted—where applicable—to group countries with the same composite score in the same year of adoption. The cumulative number of countries introducing the vaccine by year is shown in Figure S12.

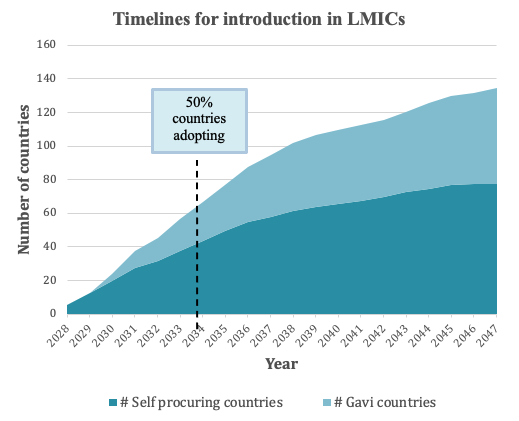

**Figure S12.**	**Assumed cumulative number of countries introducing a novel vaccine per year.**

**S1.8. Vaccine coverage targets.**

For each vaccine implementation scenario, low, medium, and high coverage targets for 5 years post-introduction were evaluated. The medium coverage target for the routine infant vaccination was 85%, based on the 2019 DTP3 (diphtheria, tetanus toxoid, and pertussis) average coverage level according to the WHO and UNICEF estimates of national immunisation coverage, with 10% uncertainty (low coverage = 75%, high coverage = 95%).^28^ Routine adolescent vaccination assumed a medium coverage target of 80% aligning with HPV coverage in South Africa combined with aggregated secondary school enrolment in China and India as assumed in Harris 2020,^29^ also with 10% uncertainty targets (low coverage = 70%, high coverage = 90%). The medium coverage target for the adolescent/adult campaign was 70% aligning with the lower bound of the MenAfriVac campaigns in sub-Saharan Africa as assumed in Harris 2020,^29^ with a wider uncertainty of 20% (low coverage = 50%, high coverage = 90%).

In the *Accelerated Scale-up* implementation, the 5-year coverage targets were achieved instantly in year 1, while in the *Basecase* and *Routine Only* implementations, the scale-up to coverage occurred linearly over 5 years.

**Appendix S2. Analysed low- and middle-income country list.**

| **Country** | **WHO Region** | **Income level^a^** | **Gavi status** | **High-TB burden^b^** | **High-TB/HIV burden^b^** | **High-MDR/RR-TB burden^b^** | **Base-case vaccine introduction year** |
| --- | --- | --- | --- | --- | --- | --- | --- |
| Afghanistan | EMR | LIC | Gavi | No | No | No | 2031 |
| Angola | AFR | LMIC | Gavi | Yes | No | Yes | 2032 |
| Albania | EUR | UMIC | Non-Gavi | No | No | No | 2035 |
| Argentina | AMR | UMIC | Non-Gavi | No | No | No | 2031 |
| Armenia | EUR | UMIC | Gavi | No | No | No | 2033 |
| Azerbaijan | EUR | UMIC | Gavi | No | No | Yes | 2028 |
| Burundi | AFR | LIC | Gavi | No | No | No | 2044 |
| Benin | AFR | LMIC | Gavi | No | No | No | 2037 |
| Burkina Faso | AFR | LIC | Gavi | No | No | No | 2039 |
| Bangladesh | SEAR | LMIC | Gavi | Yes | No | Yes | 2035 |
| Bulgaria | EUR | UMIC | Non-Gavi | No | No | No | 2029 |
| Belarus | EUR | UMIC | Non-Gavi | No | No | Yes | 2028 |
| Bolivia | AMR | LMIC | Gavi | No | No | No | 2037 |
| Brazil | AMR | UMIC | Non-Gavi | Yes | Yes | No | 2030 |
| Bhutan | SEAR | LMIC | Gavi | No | No | No | 2034 |
| Botswana | AFR | UMIC | Non-Gavi | No | Yes | No | 2028 |
| Central African Republic | AFR | LIC | Gavi | Yes | Yes | No | 2033 |
| China | WPR | UMIC | Non-Gavi | Yes | No | Yes | 2029 |
| Côte d'Ivoire | AFR | LMIC | Gavi | No | No | No | 2034 |
| Cameroon | AFR | LMIC | Gavi | No | Yes | No | 2031 |
| Colombia | AMR | UMIC | Non-Gavi | No | No | No | 2030 |
| Costa Rica | AMR | UMIC | Non-Gavi | No | No | No | 2033 |
| Cuba | AMR | UMIC | Gavi | No | No | No | 2035 |
| Dominican Republic | AMR | UMIC | Non-Gavi | No | No | No | 2031 |
| Ecuador | AMR | UMIC | Non-Gavi | No | No | No | 2033 |
| Egypt | EMR | LMIC | Non-Gavi | No | No | No | 2033 |
| Eritrea | AFR | LIC | Gavi | No | No | No | 2047 |
| Ethiopia | AFR | LIC | Gavi | Yes | Yes | No | 2030 |
| Fiji | WPR | UMIC | Non-Gavi | No | No | No | 2031 |
| Gabon | AFR | UMIC | Non-Gavi | Yes | Yes | No | 2038 |
| Georgia | EUR | UMIC | Gavi | No | No | No | 2029 |
| Ghana | AFR | LMIC | Gavi | No | No | No | 2040 |
| Guinea | AFR | LIC | Gavi | No | Yes | No | 2033 |
| Gambia | AFR | LIC | Gavi | No | No | No | 2039 |
| Equatorial Guinea | AFR | UMIC | Non-Gavi | No | No | No | 2042 |
| Guatemala | AMR | UMIC | Non-Gavi | No | No | No | 2036 |
| Honduras | AMR | LMIC | Gavi | No | No | No | 2037 |
| Indonesia | SEAR | UMIC | Gavi | Yes | Yes | Yes | 2034 |
| India | SEAR | LMIC | Gavi | Yes | Yes | Yes | 2033 |
| Iran | EMR | UMIC | Non-Gavi | No | No | No | 2031 |
| Iraq | EMR | UMIC | Non-Gavi | No | No | No | 2033 |
| Jordan | EMR | UMIC | Non-Gavi | No | No | No | 2037 |
| Kazakhstan | EUR | UMIC | Non-Gavi | No | No | Yes | 2028 |
| Kenya | AFR | LMIC | Gavi | Yes | Yes | No | 2032 |
| Kyrgyz Republic | EUR | LMIC | Gavi | No | No | Yes | 2044 |
| Cambodia | WPR | LMIC | Gavi | No | No | No | 2036 |
| Lao People’s Democratic Republic | WPR | LMIC | Gavi | No | No | No | 2035 |
| Liberia | AFR | LIC | Gavi | Yes | Yes | No | 2037 |
| Libya | EMR | UMIC | Non-Gavi | No | No | No | 2035 |
| Sri Lanka | SEAR | LMIC | Gavi | No | No | No | 2028 |
| Lesotho | AFR | LMIC | Gavi | Yes | Yes | No | 2039 |
| Morocco | EMR | LMIC | Non-Gavi | No | No | No | 2029 |
| Moldova, Republic of | EUR | LMIC | Gavi | No | No | Yes | 2034 |
| Madagascar | AFR | LIC | Gavi | No | No | No | 2031 |
| Maldives | SEAR | UMIC | Non-Gavi | No | No | No | 2034 |
| Mexico | AMR | UMIC | Non-Gavi | No | No | No | 2029 |
| Mali | AFR | LIC | Gavi | No | No | No | 2037 |
| Myanmar | SEAR | LMIC | Gavi | Yes | Yes | Yes | 2031 |
| Montenegro | EUR | UMIC | Non-Gavi | No | No | No | 2044 |
| Mongolia | WPR | LMIC | Gavi | Yes | No | Yes | 2032 |
| Mozambique | AFR | LIC | Gavi | Yes | Yes | Yes | 2032 |
| Mauritania | AFR | LMIC | Gavi | No | No | No | 2042 |
| Malawi | AFR | LIC | Gavi | No | Yes | No | 2038 |
| Malaysia | WPR | UMIC | Non-Gavi | No | No | No | 2028 |
| Namibia | AFR | UMIC | Non-Gavi | Yes | Yes | No | 2030 |
| Niger | AFR | LIC | Gavi | No | No | No | 2036 |
| Nigeria | AFR | LMIC | Gavi | Yes | Yes | Yes | 2030 |
| Nicaragua | AMR | LMIC | Gavi | No | No | No | 2047 |
| Nepal | SEAR | LMIC | Gavi | No | No | Yes | 2036 |
| Pakistan | EMR | LMIC | Gavi | Yes | No | Yes | 2031 |
| Peru | AMR | UMIC | Non-Gavi | No | No | Yes | 2029 |
| Philippines | WPR | LMIC | Non-Gavi | Yes | Yes | Yes | 2030 |
| Papua New Guinea | WPR | LMIC | Gavi | Yes | No | Yes | 2032 |
| Paraguay | AMR | UMIC | Non-Gavi | No | No | No | 2035 |
| Russian Federation | EUR | UMIC | Non-Gavi | No | Yes | Yes | 2030 |
| Rwanda | AFR | LIC | Gavi | No | No | No | 2045 |
| Sudan | EMR | LIC | Gavi | No | No | No | 2036 |
| Senegal | AFR | LMIC | Gavi | No | No | No | 2038 |
| Solomon Islands | WPR | LMIC | Gavi | No | No | No | 2047 |
| Sierra Leone | AFR | LIC | Gavi | Yes | No | No | 2037 |
| El Salvador | AMR | LMIC | Non-Gavi | No | No | No | 2039 |
| Serbia | EUR | UMIC | Non-Gavi | No | No | No | 2036 |
| South Sudan | AFR | LIC | Gavi | No | No | No | 2034 |
| São Tomé and Principe | AFR | LMIC | Gavi | No | No | No | 2044 |
| Suriname | AMR | UMIC | Non-Gavi | No | No | No | 2040 |
| Swaziland | AFR | LMIC | Non-Gavi | No | Yes | No | 2036 |
| Syrian Arab Republic | EMR | LIC | Gavi | No | No | No | 2036 |
| Chad | AFR | LIC | Gavi | No | No | No | 2033 |
| Togo | AFR | LIC | Gavi | No | No | No | 2041 |
| Thailand | SEAR | UMIC | Non-Gavi | Yes | Yes | No | 2031 |
| Tajikistan | EUR | LIC | Gavi | No | No | Yes | 2045 |
| Timor-Leste | SEAR | LMIC | Gavi | No | No | No | 2031 |
| Tunisia | EMR | LMIC | Non-Gavi | No | No | No | 2036 |
| Turkey | EUR | UMIC | Non-Gavi | No | No | No | 2030 |
| Tanzania, United Republic of | AFR | LMIC | Gavi | Yes | Yes | No | 2031 |
| Uganda | AFR | LIC | Gavi | Yes | Yes | No | 2034 |
| Ukraine | EUR | LMIC | Non-Gavi | No | No | Yes | 2033 |
| Uzbekistan | EUR | LMIC | Gavi | No | No | Yes | 2038 |
| Venezuela | AMR | UMIC | Non-Gavi | No | No | No | 2035 |
| Vietnam | WPR | LMIC | Gavi | Yes | No | Yes | 2038 |
| Vanuatu | WPR | LMIC | Non-Gavi | No | No | No | 2042 |
| Yemen | EMR | LIC | Gavi | No | No | No | 2036 |
| South Africa | AFR | UMIC | Non-Gavi | Yes | Yes | Yes | 2029 |
| Zambia | AFR | LIC | Gavi | Yes | Yes | Yes | 2034 |
| Zimbabwe | AFR | LMIC | Gavi | No | Yes | Yes | 2032 |

^a^ LIC: Gross national income (GNI) per capita of $1,085 or less; LMIC: GNI per capita of $1,086 to $4,225; UMIC: GNI per capita of $4,256 to $13,205 (World Bank 2021).

^b^ High-TB, high-TB/HIV (HIV-associated TB), and high-MDR/RR-TB (multidrug/rifampicin-resistant TB) burden countries as defined by the World Health Organization.^30^

Note: Vaccine introduction was assumed to commence in 2028 and end in 2047. See Clark et al. (https://doi.org/10.1101/2022.04.16.22273762)^1^ for introduction year methodology. AFR = African region; AMR = Region of the Americas; EMR = Eastern Mediterranean region; EUR = European region; LIC = low-income; LMIC = lower middle-income; SEAR = Southeast Asian region; TB = tuberculosis; UMIC = upper middle-income; WPR = Western Pacific region.

**Appendix S3. Sources and values of analytic inputs**

| Input | Value/Assumption/Formula | Source |
| --- | --- | --- |
| **Total costs borne by TB-affected households, per episode** | Stratified by country, income quintile, and cost category | ^31^ |
| **Fraction of TB cases incurring catastrophic costs** | Stratified by country and income quintile | ^31^ |
| **Risk ratio of TB disease across income strata** | Poorest quintile: Reference  Poorer quintile: 0·8479  Middle quintile: 0·7477  Richer quintile: 0·6221  Richest quintile: 0·5157 | Quantitative synthesis of TB prevalence surveys^9-18^ |
| **Total costs borne by TB-affected households that are not accessing care/ with untreated TB, compared to those accessing treatment** | 100% (50% and 150% in sensitivity analyses) | Assumption |
| **Discount rate** | Health outcomes: 0%  Cost outcomes: 0% (3% in sensitivity analysis) | ^32^ |
| **Concentration index** | If $z$ = area under the concentration curve, then  $C = 1 - z*2$,  where negative values indicate concentration among poor and positive values indicate concentration among rich | ^33^ |

Note: TB = tuberculosis.

**Appendix S4. Tuberculosis cases averted, costs borne by TB-affected households averted, and number of households with catastrophic costs averted by infant and adolescent/adult tuberculosis vaccines (in millions) across 105 low- and middle-income countries by income quintile.**

|  |  | Poorest | Poorer | Middle | Richer | Richest |
| --- | --- | --- | --- | --- | --- | --- |
| Tuberculosis cases averted | Infant vaccine | 2·06 (1·83–2·30) | 1·74 (1·55–1·95) | 1·18 (1·05–1·32) | 0·98 (0·87–1·10) | 0·81 (0·72–0·91) |
|  | Adolescent/adult vaccine | 9·55 (8·79–10·3) | 8·1 (7·46–8·74) | 5·6 (5·15–6·09) | 4·66 (4·28–5·07) | 3·86 (3·55–4·2) |
| Direct medical costs borne by TB-affected households averted | Infant vaccine | 207 (182–230) | 202 (179–225) | 181 (160–200) | 213 (189–235) | 237 (210–265) |
|  | Adolescent/adult vaccine | 999 (924–1082) | 984 (912–1059) | 904 (837–976) | 1076 (994–1159) | 1213 (1110–1319) |
| Direct non-medical costs borne by TB-affected households averted | Infant vaccine | 555 (495–616) | 545 (483–605) | 395 (354–440) | 393 (352–439) | 386 (346–431) |
|  | Adolescent/adult vaccine | 2589 (2402–2784) | 2544 (2362–2735) | 1895 (1756–2043) | 1888 (1749–2035) | 1854 (1710–2008) |
| Indirect costs borne by TB-affected households averted | Infant vaccine | 246 (221–271) | 440 (393–490) | 470 (423–524) | 663 (596–738) | 799 (712–896) |
|  | Adolescent/adult vaccine | 1107 (1031–1185) | 1974 (1842–2123) | 2169 (2019–2343) | 3049 (2838–3287) | 3673 (3387–3988) |
| Total costs borne by TB-affected households averted | Infant vaccine | 1007 (900–1115) | 1187 (1058–1313) | 1046 (942–1157) | 1268 (1144–1401) | 1423 (1274–1579) |
|  | Adolescent/adult vaccine | 4695 (4384–5019) | 5502 (5151–5874) | 4968 (4641–5339) | 6014 (5637–6447) | 6740 (6288–7231) |
| Number of households with catastrophic costs averted | Infant vaccine | 1·47 (1·31–1·63) | 0·97 (0·86–1·08) | 0·55 (0·49–0·61) | 0·41 (0·37–0·45) | 0·32 (0·28–0·35) |
|  | Adolescent/adult vaccine | 6·62 (6·13–7·11) | 4·26 (3·96–4·58) | 2·44 (2·26–2·64) | 1·81 (1·68–1·96) | 1·41 (1·31–1·52) |

Note: Total costs included patient direct medical, direct non-medical, and indirect costs (all undiscounted) in 2020 USD. Costs borne by TB-affected households are categorized as “catastrophic” if they exceed 20% of total household’s annual income.

**Appendix S5. Distribution of costs borne by TB-affected households averted by an adolescent/adult vaccine across all modelled strata, ordered by household income.**

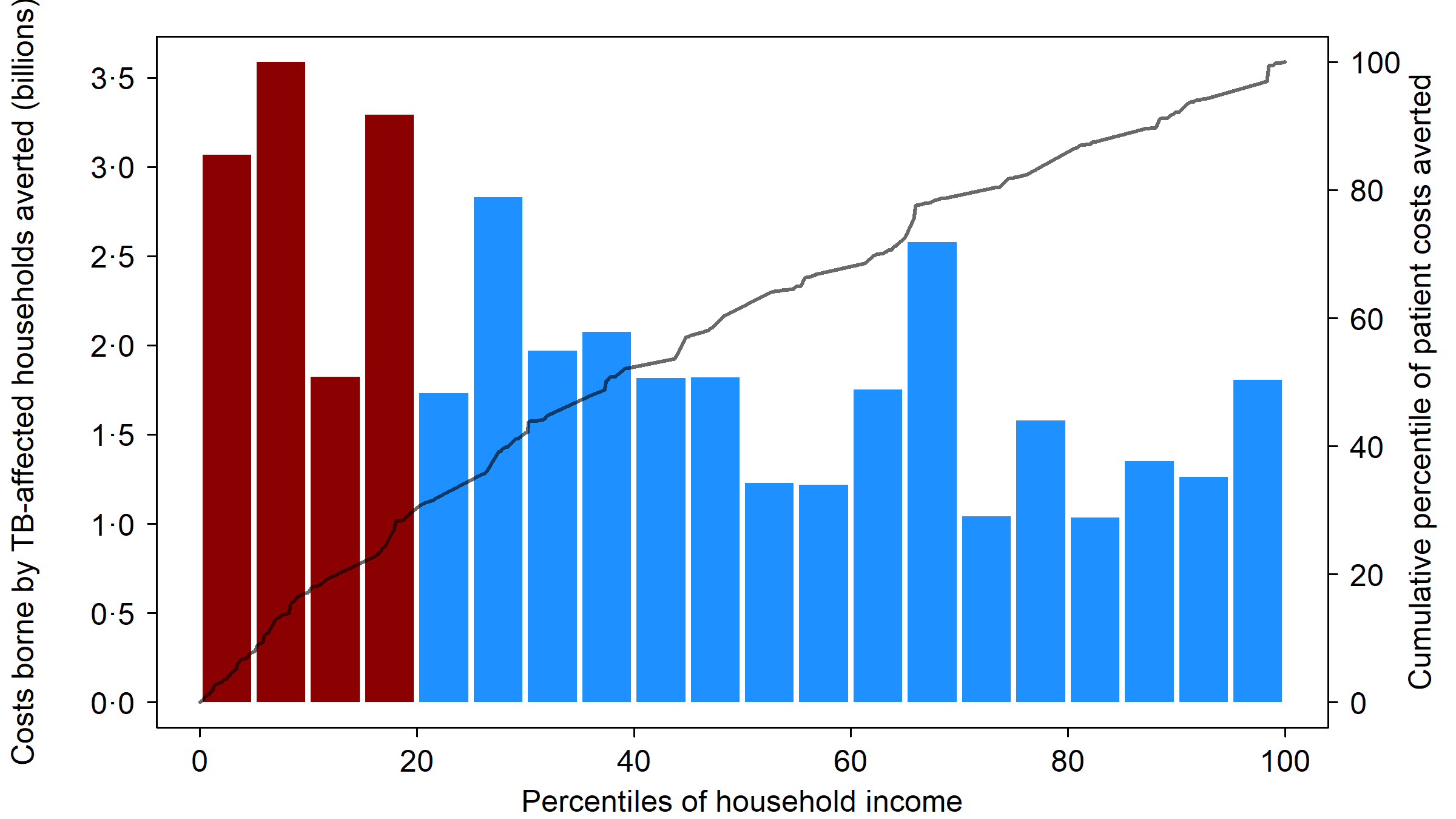

Note: TB = tuberculosis, CC = catastrophic costs. Ordering of population by household income based on average 2020 per capita GDP in purchasing power parity (PPP) dollars, for each modelled stratum (505 total strata). Bars shaded red indicate poorest 20% of modelled population by PPP GDP per capita.

**Appendix S6. Costs borne by TB-affected households averted and number of households with catastrophic costs averted by adolescent/adult vaccines (in millions): 75% efficacy scenario.**

| Country grouping | Direct medical costs borne by TB-affected households averted | Direct non-medical costs borne by TB-affected households averted | Indirect costs borne by TB-affected households averted | Total costs borne by TB-affected households averted | Number of households with catastrophic costs averted |
| --- | --- | --- | --- | --- | --- |
| All countries | 10613 (9889–11347) | 21893 (20494–23438) | 24305 (22742–26094) | 56812 (53484–60618) | 33·4 (31·2–35·8) |
| High-TB burden^a^ | 8094 (7456–8735) | 18805 (17395–20295) | 20086 (18629–21731) | 46985 (43777–50586) | 29·6 (27·3–31·8) |
| High-TB/HIV burden^a^ | 5546 (5040–6085) | 15625 (14254–17071) | 16861 (15409–18467) | 38032 (34936–41398) | 25·2 (23·1–27·4) |
| High-MDR/RR-TB burden^a^ | 8229 (7573–8945) | 17047 (15674–18476) | 18455 (16975–20092) | 43732 (40469–47176) | 26·1 (23·9–28·3) |
|  | Income level^b^ | | | | |
| LIC | 1261 (1111–1427) | 2443 (2227–2669) | 4089 (3608–4616) | 7793 (6961–8654) | 4·82 (4·39–5·25) |
| LMIC | 6023 (5454–6605) | 15437 (14085–16812) | 13720 (12541–14966) | 35180 (32139–38183) | 25·2 (23·0–27·3) |
| UMIC | 3329 (2975–3690) | 4014 (3694–4340) | 6496 (5671–7359) | 13839 (12574–15100) | 3·40 (3·04–3·79) |
|  | World region | | | | |
| AFR | 3098 (2812–3417) | 7429 (6833–8059) | 9720 (8808–10731) | 20246 (18603–22045) | 12·5 (11·6–13·4) |
| AMR | 610 (546–679) | 792 (731–853) | 762 (699–828) | 2164 (2001–2327) | 0·45 (0·42–0·48) |
| EMR | 1212 (1026–1413) | 2029 (1693–2384) | 3080 (2574–3632) | 6320 (5351–7363) | 3·39 (2·78–4·06) |
| EUR | 631 (536–746) | 326 (291–361) | 443 (388–502) | 1400 (1231–1592) | 0·22 (0·20–0·25) |
| SEAR | 2454 (2061–2892) | 8189 (7032–9432) | 6323 (5374–7332) | 16966 (14500–19571) | 13·0 (11·2–14·9) |
| WPR | 2609 (2304–2935) | 3129 (2830–3465) | 3978 (3509–4510) | 9715 (8756–10750) | 3·90 (3·38–4·52) |

Note: Values in parentheses represent equal-tailed 95% credible intervals. Total costs included patient direct medical, direct non-medical, and indirect costs (all undiscounted) in 2020 USD. Costs borne by TB-affected households are categorized as “catastrophic” if they exceed 20% of total household’s annual income.

^a^ High-TB, high-TB/HIV (HIV-associated TB), and high-MDR/RR-TB (multidrug/rifampicin-resistant TB) burden countries as defined by the World Health Organization.

^b^ LIC: Gross national income (GNI) per capita of $1,085 or less; LMIC: GNI per capita of $1,086 to $4,225; UMIC: GNI per capita of $4,256 to $13,205 (World Bank 2021).

Note: All countries include 105 low- and middle-income countries analysed. AFR = African region; AMR = Region of the Americas; EMR = Eastern Mediterranean region; EUR = European region; GDP = gross domestic product; LIC = low-income; LMIC = lower middle-income; SEAR = Southeast Asian region; UMIC = upper middle-income; WPR = Western Pacific region.

**Appendix S7. Costs borne by TB-affected households averted and number of households with catastrophic costs averted by infant vaccines (in millions): accelerated scale-up scenario.**

| Country grouping | Direct medical costs borne by TB-affected households averted | Direct non-medical costs borne by TB-affected households averted | Indirect costs borne by TB-affected households averted | Total costs borne by TB-affected households averted | Number of households with catastrophic costs averted |
| --- | --- | --- | --- | --- | --- |
| All countries | 2441 (2190–2700) | 5434 (4883–6053) | 5972 (5397–6629) | 13847 (12506–15274) | 8·83 (7·93–9·80) |
| High-TB burden^a^ | 1800 (1571–2035) | 4639 (4098–5253) | 4717 (4213–5285) | 11156 (9922–12518) | 7·73 (6·87–8·70) |
| High-TB/HIV burden^a^ | 1484 (1260–1708) | 4059 (3544–4647) | 4123 (3637–4678) | 9666 (8494–11002) | 6·66 (5·84–7·58) |
| High-MDR/RR-TB burden^a^ | 1794 (1561–2036) | 4179 (3655–4783) | 4241 (3751–4803) | 10214 (8997–11559) | 6·77 (5·95–7·72) |
|  | Income level^b^ | | | | |
| LIC | 449 (372–532) | 807 (704–920) | 1456 (1228–1721) | 2712 (2308–3148) | 1·54 (1·34–1·79) |
| LMIC | 1686 (1461–1928) | 4223 (3693–4836) | 3708 (3263–4252) | 9616 (8443–11011) | 6·87 (6·04–7·85) |
| UMIC | 306 (267–348) | 405 (337–478) | 808 (612–1042) | 1519 (1229–1850) | 0·42 (0·32–0·52) |
|  | World region | | | | |
| AFR | 1016 (862–1159) | 2290 (2001–2581) | 2614 (2297–2951) | 5919 (5199–6649) | 3·81 (3·37–4·28) |
| AMR | 64·2 (55·8–72·5) | 65·7 (59·8–73·1) | 67·6 (60·8–75·3) | 198 (179–218) | 0·039 (0·035–0·044) |
| EMR | 404 (316–492) | 636 (494–791) | 1071 (820–1313) | 2111 (1649–2578) | 1·06 (0·80–1·35) |
| EUR | 122 (95·0–156) | 46·3 (37·5–57·4) | 75·7 (60·8–94·6) | 243 (196–307) | 0·04 (0·03–0·05) |
| SEAR | 575 (430–755) | 1955 (1536–2477) | 1531 (1195–1942) | 4061 (3173–5163) | 3·12 (2·47–3·90) |
| WPR | 261 (219–310) | 441 (361–527) | 612 (484–747) | 1314 (1075–1558) | 0·76 (0·59–0·93) |

Note: Values in parentheses represent equal-tailed 95% credible intervals. Total costs included patient direct medical, direct non-medical, and indirect costs (all undiscounted) in 2020 USD. Costs borne by TB-affected households are categorized as “catastrophic” if they exceed 20% of total household’s annual income.

^a^ High-TB, high-TB/HIV (HIV-associated TB), and high-MDR/RR-TB (multidrug/rifampicin-resistant TB) burden countries as defined by the World Health Organization.

^b^ LIC: Gross national income (GNI) per capita of $1,085 or less; LMIC: GNI per capita of $1,086 to $4,225; UMIC: GNI per capita of $4,256 to $13,205 (World Bank 2021).

Note: All countries include 105 low- and middle-income countries analysed. AFR = African region; AMR = Region of the Americas; EMR = Eastern Mediterranean region; EUR = European region; GDP = gross domestic product; LIC = low-income; LMIC = lower middle-income; SEAR = Southeast Asian region; UMIC = upper middle-income; WPR = Western Pacific region.

**Appendix S8. Costs borne by TB-affected households averted and number of households with catastrophic costs averted by adolescent/adult tuberculosis vaccines (in millions): accelerated scale-up scenario.**

| Country grouping | Direct medical costs borne by TB-affected households averted | Direct non-medical costs borne by TB-affected households averted | Indirect costs borne by TB-affected households averted | Total costs borne by TB-affected households averted | Number of households with catastrophic costs averted |
| --- | --- | --- | --- | --- | --- |
| All countries | 11021 (10308–11771) | 22358 (20909–23896) | 24279 (22813–25976) | 57658 (54410–61314) | 33·9 (31·7–36·3) |
| High-TB burden^a^ | 8015 (7389–8634) | 18794 (17329–20281) | 19566 (18171–21086) | 46375 (43173–49852) | 29·5 (27·2–31·8) |
| High-TB/HIV burden^a^ | 5505 (4986–6025) | 15672 (14328–17104) | 16345 (15020–17807) | 37522 (34506–40766) | 25·1 (23–27·4) |
| High-MDR/RR-TB burden^a^ | 8430 (7776–9130) | 17210 (15762–18693) | 18068 (16691–19547) | 43708 (40371–47120) | 26·3 (24·0–28·6) |
|  | Income level^b^ | | | | |
| LIC | 1352 (1197–1524) | 2579 (2369–2809) | 4263 (3790–4788) | 8194 (7370–9073) | 4·97 (4·56–5·38) |
| LMIC | 6537 (5976–7124) | 16118 (14739–17531) | 14291 (13060–15559) | 36947 (33837–40094) | 26·0 (23·8–28·2) |
| UMIC | 3132 (2825–3482) | 3660 (3401–3943) | 5725 (5058–6420) | 12517 (11464–13560) | 3·00 (2·71–3·31) |
|  | World region | | | | |
| AFR | 3042 (2796–3296) | 7198 (6706–7706) | 9173 (8400–9995) | 19413 (17988–20873) | 12·0 (11·3–12·8) |
| AMR | 648 (580–718) | 825 (767–885) | 794 (733–859) | 2267 (2104–2425) | 0·48 (0·44–0·51) |
| EMR | 1197 (1020–1390) | 1986 (1667–2305) | 3013 (2535–3526) | 6196 (5290–7196) | 3·28 (2·71–3·89) |
| EUR | 909 (774–1073) | 415 (373–460) | 609 (535–689) | 1933 (1713–2190) | 0·31 (0·28–0·35) |
| SEAR | 2669 (2238–3114) | 8974 (7684–10314) | 6888 (5852–7939) | 18531 (15779–21289) | 14·2 (12·3–16·2) |
| WPR | 2556 (2268–2879) | 2961 (2707–3249) | 3801 (3386–4258) | 9318 (8475–10274) | 3·64 (3·22–4·17) |

Note: Values in parentheses represent equal-tailed 95% credible intervals. Total costs included patient direct medical, direct non-medical, and indirect costs (all undiscounted) in 2020 USD. Costs borne by TB-affected households are categorized as “catastrophic” if they exceed 20% of total household’s annual income.

^a^ High-TB, high-TB/HIV (HIV-associated TB), and high-MDR/RR-TB (multidrug/rifampicin-resistant TB) burden countries as defined by the World Health Organization.

^b^ LIC: Gross national income (GNI) per capita of $1,025 or less; LMIC: GNI per capita of $1,026 to $3,995; UMIC: GNI per capita of $3,996 to $12,375 (World Bank 2019).

Note: All countries include 105 low- and middle-income countries analysed. AFR = African region; AMR = Region of the Americas; EMR = Eastern Mediterranean region; EUR = European region; GDP = gross domestic product; LIC = low-income; LMIC = lower middle-income; SEAR = Southeast Asian region; UMIC = upper middle-income; WPR = Western Pacific region.

**Appendix S9. Costs borne by TB-affected households averted and number of households with catastrophic costs averted by infant tuberculosis vaccines (in millions), assuming costs for untreated cases are 0.5x treated cases.**

| Country grouping | Direct medical costs borne by TB-affected households averted | Direct non-medical costs borne by TB-affected households averted | Indirect costs borne by TB-affected households averted | Total costs borne by TB-affected households averted | Number of households with catastrophic costs averted |
| --- | --- | --- | --- | --- | --- |
| All countries | 713 (641–781) | 1570 (1422–1718) | 1885 (1713–2073) | 4168 (3798–4572) | 2·80 (2·53–3·07) |
| High-TB burden^a^ | 546 (481–609) | 1371 (1228–1522) | 1504 (1345–1675) | 3422 (3075–3784) | 2·49 (2·22–2·76) |
| High-TB/HIV burden^a^ | 441 (379–501) | 1174 (1037–1319) | 1305 (1161–1467) | 2920 (2589–3280) | 2·12 (1·88–2·36) |
| High-MDR/RR-TB burden^a^ | 525 (460–588) | 1223 (1080–1377) | 1360 (1208–1531) | 3108 (2770–3474) | 2·16 (1·91–2·42) |
|  | Income level^b^ | | | | |
| LIC | 136 (110–165) | 234 (202–271) | 459 (379–553) | 829 (694–982) | 0·51 (0·44–0·60) |
| LMIC | 464 (401–527) | 1180 (1038–1337) | 1092 (966–1238) | 2736 (2416–3096) | 2·12 (1·87–2·38) |
| UMIC | 113 (98–129) | 156 (130–185) | 335 (255–426) | 603 (489–734) | 0·18 (0·14–0·23) |
|  | World region | | | | |
| AFR | 300 (252–346) | 688 (599–783) | 835 (721–951) | 1822 (1588–2062) | 1·31 (1·15–1·48) |
| AMR | 21·8 (19·0–24·7) | 20·5 (18·5–22·5) | 22·7 (20·4–25·1) | 65 (58·6–71·4) | 0·012 (0·011–0·013) |
| EMR | 146 (115–178) | 230 (181–285) | 402 (316–490) | 778 (618–948) | 0·42 (0·32–0·54) |
| EUR | 16·7 (13·8–20·1) | 8·14 (7·13–9·16) | 11·6 (9·90–13·4) | 36·5 (31·2–42·1) | 0·006 (0·005–0·007) |
| SEAR | 141 (109–178) | 463 (368–570) | 386 (312–477) | 990 (789–1222) | 0·77 (0·62–0·94) |
| WPR | 87·7 (74·6–102) | 161 (130–192) | 228 (179–279) | 477 (385–572) | 0·29 (0·22–0·36) |

Note: Values in parentheses represent equal-tailed 95% credible intervals. Total costs included patient direct medical, direct non-medical, and indirect costs (all undiscounted) in 2020 USD. Costs borne by TB-affected households are categorized as “catastrophic” if they exceed 20% of total household’s annual income.

^a^ High-TB, high-TB/HIV (HIV-associated TB), and high-MDR/RR-TB (multidrug/rifampicin-resistant TB) burden countries as defined by the World Health Organization.

^b^ LIC: Gross national income (GNI) per capita of $1,085 or less; LMIC: GNI per capita of $1,086 to $4,225; UMIC: GNI per capita of $4,256 to $13,205 (World Bank 2021).

Note: All countries include 105 low- and middle-income countries analysed. AFR = African region; AMR = Region of the Americas; EMR = Eastern Mediterranean region; EUR = European region; GDP = gross domestic product; LIC = low-income; LMIC = lower middle-income; SEAR = Southeast Asian region; UMIC = upper middle-income; WPR = Western Pacific region.

**Appendix S10. Costs borne by TB-affected households averted and number of households with catastrophic costs averted by adolescent/adult tuberculosis vaccines (in millions), assuming costs for untreated cases are 0.5x treated cases.**

| Country grouping | Direct medical costs borne by TB-affected households averted | Direct non-medical costs borne by TB-affected households averted | Indirect costs borne by TB-affected households averted | Total costs borne by TB-affected households averted | Number of households with catastrophic costs averted |
| --- | --- | --- | --- | --- | --- |
| All countries | 5727 (5360–6096) | 11734 (11011–12476) | 13356 (12584–14211) | 30817 (29181–32592) | 18·7 (17·6–20·0) |
| High-TB burden^a^ | 4313 (3988–4636) | 10067 (9358–10777) | 10957 (10241–11712) | 25337 (23755–26997) | 16·5 (15·3–17·7) |
| High-TB/HIV burden^a^ | 2894 (2644–3141) | 8281 (7606–8967) | 9118 (8426–9856) | 20293 (18746–21929) | 14·0 (12·9–15·1) |
| High-MDR/RR-TB burden^a^ | 4405 (4075–4750) | 9119 (8416–9817) | 10073 (9374–10810) | 23598 (22050–25247) | 14·5 (13·4–15·7) |
|  | Income level^b^ | | | | |
| LIC | 705 (625–794) | 1339 (1229–1458) | 2323 (2064–2617) | 4367 (3924–4826) | 2·85 (2·60–3·09) |
| LMIC | 3163 (2899–3427) | 8197 (7537–8902) | 7480 (6863–8106) | 18841 (17370–20375) | 14·0 (12·8–15·1) |
| UMIC | 1859 (1677–2053) | 2197 (2049–2349) | 3554 (3172–3950) | 7610 (6999–8190) | 1·90 (1·71–2·09) |
|  | World region | | | | |
| AFR | 1496 (1383–1613) | 3651 (3418–3893) | 5000 (4602–5453) | 10148 (9466–10864) | 6·77 (6·34–7·21) |
| AMR | 361 (324–402) | 457 (424–491) | 451 (416–489) | 1269 (1179–1359) | 0·27 (0·25–0·29) |
| EMR | 687 (589–796) | 1143 (969–1329) | 1797 (1533–2096) | 3626 (3114–4177) | 2·02 (1·68–2·37) |
| EUR | 363 (312–422) | 187 (169–204) | 258 (229–289) | 807 (719–907) | 0·13 (0·12–0·15) |
| SEAR | 1371 (1165–1583) | 4555 (3948–5185) | 3595 (3080–4114) | 9522 (8178–10892) | 7·33 (6·36–8·32) |
| WPR | 1450 (1291–1609) | 1740 (1594–1893) | 2255 (2021–2498) | 5445 (4966–5939) | 2·21 (1·95–2·49) |

Note: Values in parentheses represent equal-tailed 95% credible intervals. Total costs included patient direct medical, direct non-medical, and indirect costs (all undiscounted) in 2020 USD. Costs borne by TB-affected households are categorized as “catastrophic” if they exceed 20% of total household’s annual income.

^a^ High-TB, high-TB/HIV (HIV-associated TB), and high-MDR/RR-TB (multidrug/rifampicin-resistant TB) burden countries as defined by the World Health Organization.

^b^ LIC: Gross national income (GNI) per capita of $1,085 or less; LMIC: GNI per capita of $1,086 to $4,225; UMIC: GNI per capita of $4,256 to $13,205 (World Bank 2021).

Note: All countries include 105 low- and middle-income countries analysed. AFR = African region; AMR = Region of the Americas; EMR = Eastern Mediterranean region; EUR = European region; GDP = gross domestic product; LIC = low-income; LMIC = lower middle-income; SEAR = Southeast Asian region; UMIC = upper middle-income; WPR = Western Pacific region.

**Appendix S11. Total costs borne by TB-affected households averted in the base-case (Panel A) and assuming costs for untreated cases are 0.5x treated cases (Panel B) by within-country income quintile comparing infant vaccine to adolescent/adult vaccine.**

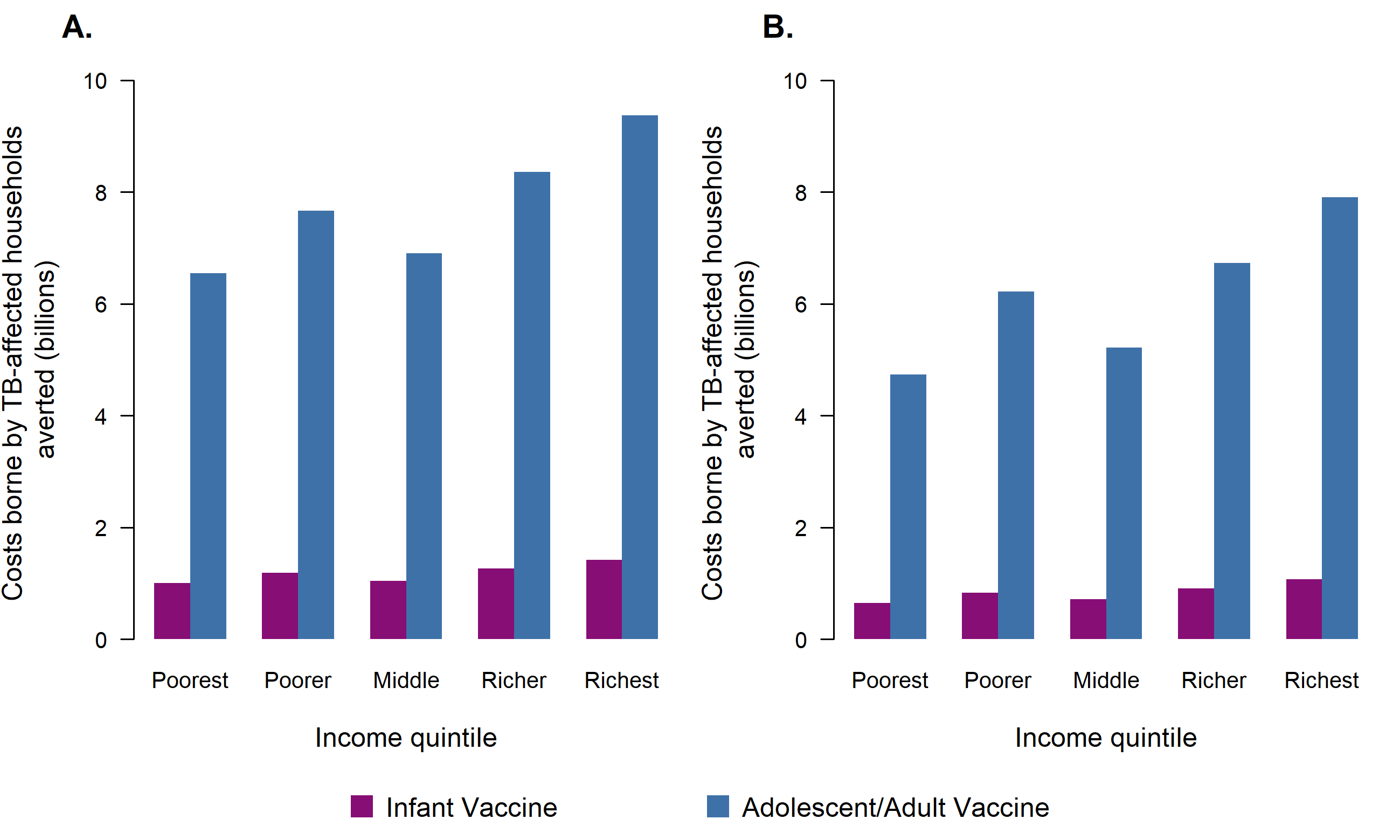

Note: Total costs included patient direct medical, direct non-medical, and indirect costs (all undiscounted) in 2020 USD.

**Appendix S12. Costs borne by TB-affected households averted and number of households with catastrophic costs averted by infant tuberculosis vaccines (in millions), assuming costs for untreated cases are 1.5x treated cases.**

| Country grouping | Direct medical costs borne by TB-affected households averted | Direct non-medical costs borne by TB-affected households averted | Indirect costs borne by TB-affected households averted | Total costs borne by TB-affected households averted | Number of households with catastrophic costs averted |
| --- | --- | --- | --- | --- | --- |
| All countries | 1359 (1199–1504) | 2958 (2629–3296) | 3329 (2984–3712) | 7646 (6855–8491) | 4·21 (3·75–4·68) |
| High-TB burden^a^ | 1082 (934–1218) | 2612 (2292–2949) | 2720 (2391–3071) | 6413 (5651–7216) | 3·80 (3·35–4·27) |
| High-TB/HIV burden^a^ | 896 (752–1027) | 2282 (1979–2607) | 2391 (2081–2734) | 5569 (4807–6346) | 3·26 (2·85–3·70) |
| High-MDR/RR-TB burden^a^ | 1041 (895–1177) | 2343 (2030–2678) | 2465 (2147–2815) | 5850 (5120–6649) | 3·32 (2·89–3·77) |
|  | Income level^b^ | | | | |
| LIC | 225 (180–274) | 399 (337–467) | 739 (596–895) | 1364 (1119–1628) | 0·66 (0·57–0·79) |
| LMIC | 934 (794–1071) | 2282 (1974–2615) | 2016 (1752–2322) | 5232 (4526–5995) | 3·29 (2·87–3·76) |
| UMIC | 199 (170–230) | 278 (221–340) | 574 (413–773) | 1051 (813–1339) | 0·26 (0·19–0·33) |
|  | World region | | | | |
| AFR | 667 (546–770) | 1469 (1251–1674) | 1641 (1391–1895) | 3777 (3221–4300) | 2·03 (1·77–2·30) |
| AMR | 32·2 (27·7–37·0) | 31·2 (27·5–35·0) | 33·2 (29·4–37·5) | 96·6 (85·9–108) | 0·016 (0·014–0·018) |
| EMR | 235 (179–291) | 375 (282–472) | 624 (466–776) | 1234 (935–1530) | 0·55 (0·40–0·72) |
| EUR | 27·0 (21·4–33·3) | 12·9 (10·8–15·0) | 18·1 (14·8–21·7) | 58·0 (47·3–69·7) | 0·008 (0·007–0·010) |
| SEAR | 241 (178–322) | 798 (610–1028) | 640 (497–831) | 1679 (1286–2181) | 1·17 (0·91–1·49) |
| WPR | 157 (131–187) | 272 (214–336) | 372 (282–469) | 802 (637–982) | 0·43 (0·31–0·54) |

Note: Values in parentheses represent equal-tailed 95% credible intervals. Total costs included patient direct medical, direct non-medical, and indirect costs (all undiscounted) in 2020 USD. Costs borne by TB-affected households are categorized as “catastrophic” if they exceed 20% of total household’s annual income.

^a^ High-TB, high-TB/HIV (HIV-associated TB), and high-MDR/RR-TB (multidrug/rifampicin-resistant TB) burden countries as defined by the World Health Organization.

^b^ LIC: Gross national income (GNI) per capita of $1,085 or less; LMIC: GNI per capita of $1,086 to $4,225; UMIC: GNI per capita of $4,256 to $13,205 (World Bank 2021).

Note: All countries include 105 low- and middle-income countries analysed. AFR = African region; AMR = Region of the Americas; EMR = Eastern Mediterranean region; EUR = European region; GDP = gross domestic product; LIC = low-income; LMIC = lower middle-income; SEAR = Southeast Asian region; UMIC = upper middle-income; WPR = Western Pacific region.

**Appendix S13. Costs borne by TB-affected households averted and number of households with catastrophic costs averted by adolescent/adult tuberculosis vaccines (in millions), assuming costs for untreated cases are 1.5x treated cases.**

| Country grouping | Direct medical costs borne by TB-affected households averted | Direct non-medical costs borne by TB-affected households averted | Indirect costs borne by TB-affected households averted | Total costs borne by TB-affected households averted | Number of households with catastrophic costs averted |
| --- | --- | --- | --- | --- | --- |
| All countries | 8777 (8141–9446) | 18241 (16978–19648) | 19883 (18497–21500) | 46902 (43898–50346) | 25·4 (23·6–27·2) |
| High-TB burden^a^ | 6734 (6155–7304) | 15691 (14430–17061) | 16445 (15154–17918) | 38869 (35905–42123) | 22·5 (20·7–24·3) |
| High-TB/HIV burden^a^ | 4720 (4260–5225) | 13158 (11948–14530) | 13896 (12627–15343) | 31774 (28970–34823) | 19·2 (17·5–21·0) |
| High-MDR/RR-TB burden^a^ | 6823 (6243–7448) | 14232 (12967–15608) | 15102 (13793–16555) | 36158 (33232–39377) | 19·9 (18·1–21·7) |
|  | Income level^b^ | | | | |
| LIC | 1048 (913–1192) | 2033 (1831–2243) | 3362 (2924–3832) | 6443 (5705–7218) | 3·55 (3·22–3·89) |
| LMIC | 5128 (4612–5664) | 13059 (11839–14361) | 11418 (10374–12493) | 29605 (26902–32463) | 19·3 (17·6–21·1) |
| UMIC | 2601 (2323–2904) | 3149 (2867–3444) | 5104 (4373–5891) | 10854 (9736–11996) | 2·47 (2·19–2·76) |
|  | World region | | | | |
| AFR | 2756 (2485–3067) | 6517 (5951–7120) | 8194 (7342–9132) | 17468 (15914–19164) | 9·46 (8·77–10·2) |
| AMR | 468 (416–524) | 604 (555–655) | 576 (527–627) | 1648 (1513–1782) | 0·32 (0·30–0·35) |
| EMR | 1006 (839–1187) | 1678 (1371–1998) | 2526 (2063–3016) | 5210 (4315–6144) | 2·53 (2·05–3·08) |
| EUR | 494 (414–590) | 252 (223–282) | 342 (294–393) | 1087 (940–1249) | 0·16 (0·14–0·18) |
| SEAR | 2006 (1650–2415) | 6721 (5684–7838) | 5130 (4295–6059) | 13856 (11616–16292) | 9·97 (8·48–11·5) |
| WPR | 2047 (1789–2334) | 2469 (2208–2760) | 3116 (2715–3566) | 7632 (6820–8553) | 2·90 (2·50–3·39) |

Note: Values in parentheses represent equal-tailed 95% credible intervals. Total costs included patient direct medical, direct non-medical, and indirect costs (all undiscounted) in 2020 USD. Costs borne by TB-affected households are categorized as “catastrophic” if they exceed 20% of total household’s annual income.

^a^ High-TB, high-TB/HIV (HIV-associated TB), and high-MDR/RR-TB (multidrug/rifampicin-resistant TB) burden countries as defined by the World Health Organization.

^b^ LIC: Gross national income (GNI) per capita of $1,085 or less; LMIC: GNI per capita of $1,086 to $4,225; UMIC: GNI per capita of $4,256 to $13,205 (World Bank 2021).

Note: All countries include 105 low- and middle-income countries analysed. AFR = African region; AMR = Region of the Americas; EMR = Eastern Mediterranean region; EUR = European region; GDP = gross domestic product; LIC = low-income; LMIC = lower middle-income; SEAR = Southeast Asian region; UMIC = upper middle-income; WPR = Western Pacific region.

**Appendix S14. Total costs borne by TB-affected households averted in the base-case (Panel A) and assuming costs for untreated cases are 1.5x treated cases (Panel B) by within-country income quintile comparing infant vaccine to adolescent/adult vaccine.**

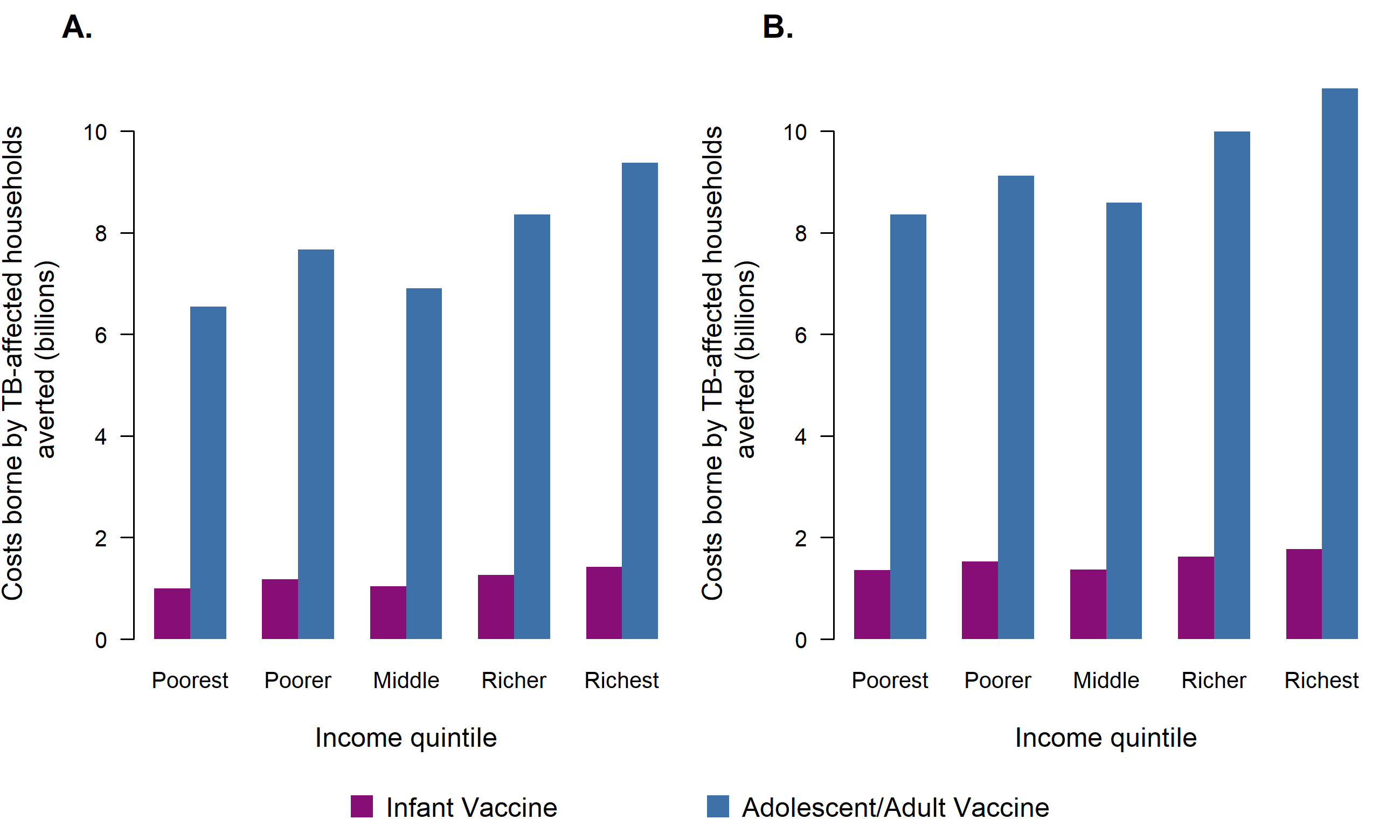

Note: Total costs included patient direct medical, direct non-medical, and indirect costs (all undiscounted) in 2020 USD.

**Appendix S15. Number of households with catastrophic costs averted in the base-case (Panel A) and assuming costs for untreated cases are 0.5x treated cases (Panel B) by within-country income quintile comparing infant vaccine to adolescent/adult vaccine.**

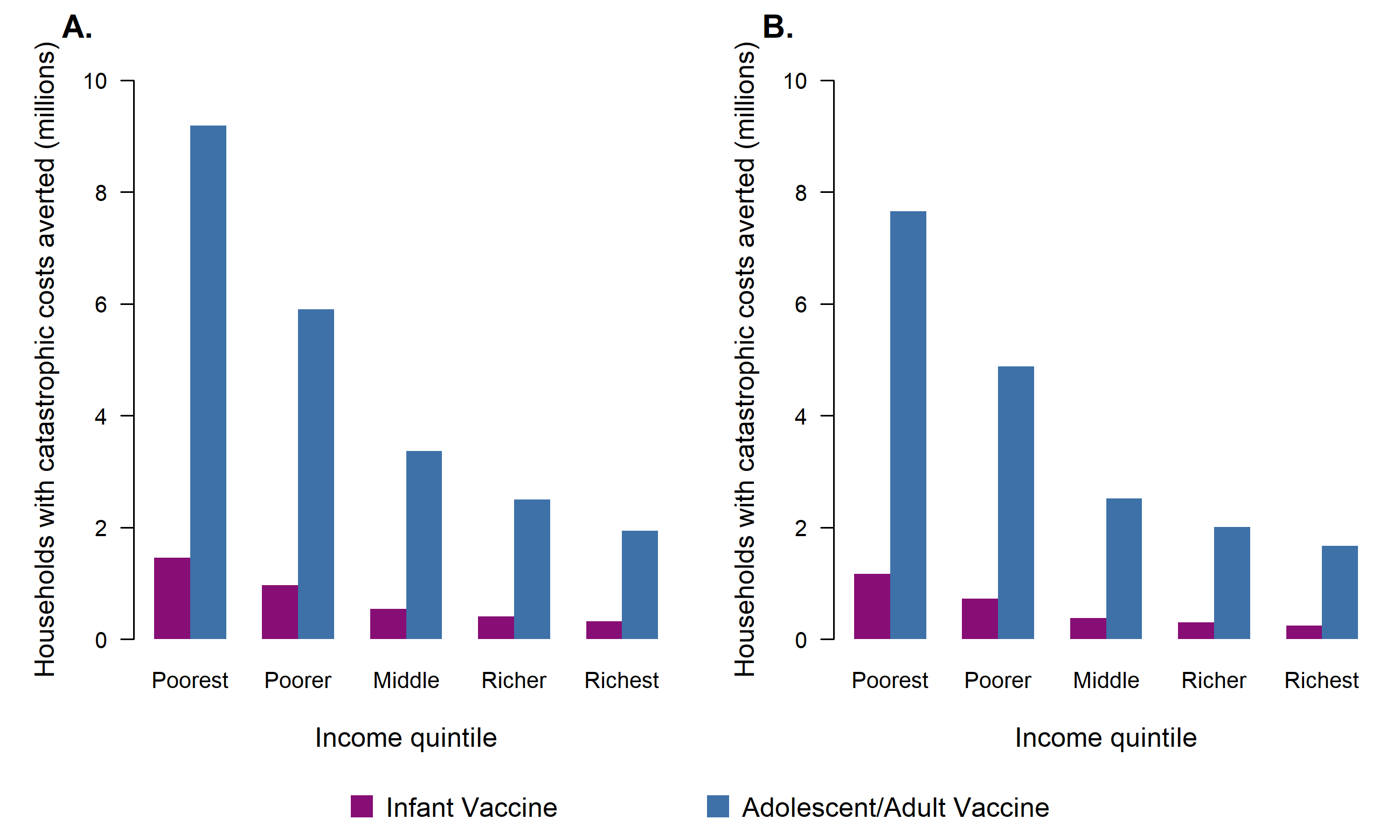

Note: Catastrophic costs included patient direct medical, direct non-medical, and indirect costs. Catastrophic costs are defined as instances where the patient costs incurred during an episode of TB disease exceed 20% of total annual household income.

**Appendix S16. Number of households with catastrophic costs averted in the base-case (Panel A) and assuming costs for untreated cases are 1.5x treated cases (Panel B) by within-country income quintile comparing infant vaccine to adolescent/adult vaccine.**

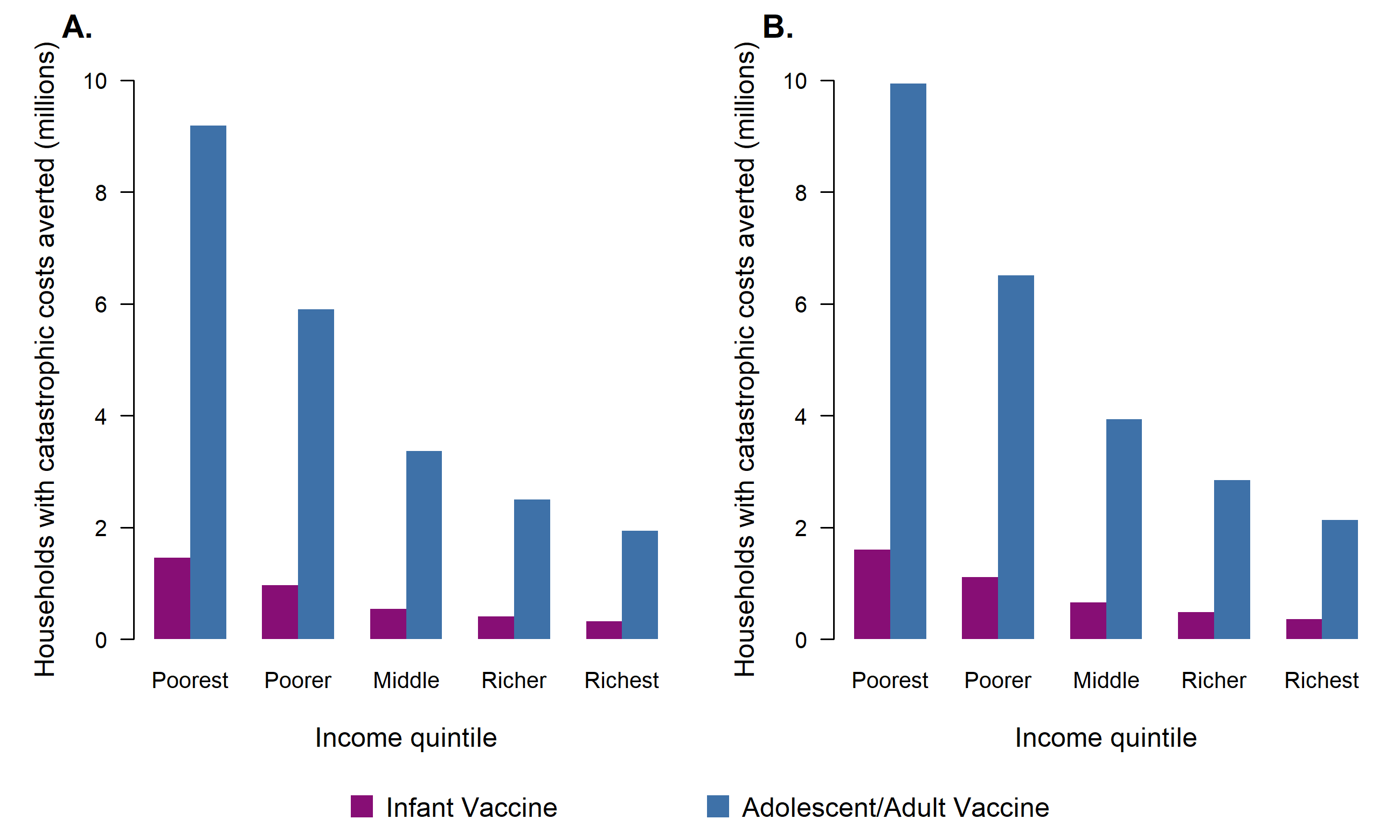

Note: Catastrophic costs included patient direct medical, direct non-medical, and indirect costs. Catastrophic costs are defined as instances where the patient costs incurred during an episode of TB disease exceed 20% of total annual household income.

**Appendix S17. Number of households with catastrophic costs averted by infant tuberculosis vaccines (in millions), assuming thresholds of 10%, 20% (base-case), and 25%.**

| Country grouping | 10% threshold | | 20% threshold | | 25% threshold | |
| --- | --- | --- | --- | --- | --- | --- |
|  | Number of households with catastrophic direct medical costs averted | Number of households with catastrophic costs averted | Number of households with catastrophic direct medical costs averted | Number of households with catastrophic costs averted | Number of households with catastrophic direct medical costs averted | Number of households with catastrophic costs averted |
| All countries | 1·15 (1·02–1·28) | 5·10 (4·56–5·67) | 0·63 (0·56–0·70) | 3·69 (3·31–4·08) | 0·51 (0·45–0·57) | 3·20 (2·87–3·54) |
| High-TB burden^a^ | 1·00 (0·87–1·12) | 4·64 (4·11–5·21) | 0·53 (0·46–0·60) | 3·32 (2·94–3·70) | 0·43 (0·37–0·48) | 2·86 (2·53–3·19) |
| High-TB/HIV burden^a^ | 0·85 (0·73–0·97) | 3·94 (3·46–4·48) | 0·46 (0·39–0·52) | 2·84 (2·50–3·21) | 0·37 (0·31–0·42) | 2·46 (2·17–2·77) |
| High-MDR/RR-TB burden^a^ | 0·90 (0·77–1·02) | 4·06 (3·54–4·61) | 0·48 (0·41–0·55) | 2·89 (2·53–3·27) | 0·39 (0·33–0·44) | 2·49 (2·18–2·81) |
|  | Income level^b^ | | | | | |
| LIC | 0·20 (0·17–0·23) | 0·76 (0·65–0·90) | 0·12 (0·10–0·14) | 0·61 (0·52–0·73) | 0·10 (0·08–0·11) | 0·55 (0·47–0·66) |
| LMIC | 0·89 (0·76–1·01) | 4·03 (3·52–4·60) | 0·48 (0·41–0·54) | 2·85 (2·50–3·25) | 0·38 (0·33–0·44) | 2·45 (2·15–2·79) |
| UMIC | 0·07 (0·05–0·08) | 0·31 (0·23–0·39) | 0·04 (0·03–0·04) | 0·23 (0·17–0·29) | 0·03 (0·02–0·04) | 0·20 (0·15–0·26) |
|  | World region | | | | | |
| AFR | 0·58 (0·50–0·67) | 2·36 (2·05–2·67) | 0·32 (0·27–0·37) | 1·78 (1·55–2·01) | 0·26 (0·22–0·30) | 1·56 (1·36–1·77) |
| AMR | 0·006 (0·005–0·007) | 0·020 (0·018–0·023) | 0·004 (0·003–0·004) | 0·015 (0·013–0·016) | 0·0030 (0·0026–0·0034) | 0·013 (0·011–0·014) |
| EMR | 0·19 (0·15–0·24) | 0·66 (0·48–0·86) | 0·11 (0·09–0·14) | 0·51 (0·38–0·65) | 0·09 (0·07–0·12) | 0·45 (0·34–0·58) |
| EUR | 0·005 (0·004–0·006) | 0·011 (0·009–0·013) | 0·003 (0·003–0·004) | 0·008 (0·006–0·009) | 0·003 (0·002–0·003) | 0·007 (0·005–0·008) |
| SEAR | 0·27 (0·21–0·35) | 1·50 (1·17–1·91) | 0·14 (0·11–0·18) | 1·01 (0·80–1·28) | 0·11 (0·08–0·14) | 0·86 (0·68–1·08) |
| WPR | 0·09 (0·07–0·12) | 0·55 (0·41–0·69) | 0·05 (0·04–0·06) | 0·37 (0·28–0·47) | 0·04 (0·03–0·05) | 0·32 (0·24–0·40) |

Note: Values in parentheses represent equal-tailed 95% credible intervals. Catastrophic costs are defined as instances where the patient costs (either direct medical only or total) incurred during an episode of TB disease exceed either 10%, 20%, or 25% of total annual household income.

^a^ High-TB, high-TB/HIV (HIV-associated TB), and high-MDR/RR-TB (multidrug/rifampicin-resistant TB) burden countries as defined by the World Health Organization.

^b^ LIC: Gross national income (GNI) per capita of $1,085 or less; LMIC: GNI per capita of $1,086 to $4,225; UMIC: GNI per capita of $4,256 to $13,205 (World Bank 2021).

Note: All countries include 105 low- and middle-income countries analysed. AFR = African region; AMR = Region of the Americas; EMR = Eastern Mediterranean region; EUR = European region; GDP = gross domestic product; LIC = low-income; LMIC = lower middle-income; SEAR = Southeast Asian region; UMIC = upper middle-income; WPR = Western Pacific region.

**Appendix S18. Number of households with catastrophic costs averted by adolescent/adult tuberculosis vaccines (in millions), assuming thresholds of 10%, 20% (base-case), and 25%.**

| Country grouping | 10% threshold | | 20% threshold | | 25% threshold | |
| --- | --- | --- | --- | --- | --- | --- |
|  | Number of households with catastrophic direct medical costs averted | Number of households with catastrophic costs averted | Number of households with catastrophic direct medical costs averted | Number of households with catastrophic costs averted | Number of households with catastrophic direct medical costs averted | Number of households with catastrophic costs averted |
| All countries | 6·97 (6·43–7·52) | 32·3 (30·0–34·6) | 3·75 (3·46–4·05) | 22·9 (21·4–24·5) | 3·01 (2·78–3·25) | 19·7 (18·4–21·1) |
| High-TB burden^a^ | 5·91 (5·40–6·43) | 28·9 (26·7–31·2) | 3·12 (2·85–3·40) | 20·2 (18·7–21·8) | 2·48 (2·27–2·71) | 17·4 (16·0–18·7) |
| High-TB/HIV burden^a^ | 4·89 (4·43–5·36) | 24·4 (22·3–26·7) | 2·57 (2·32–2·84) | 17·3 (15·8–18·8) | 2·05 (1·84–2·26) | 14·8 (13·6–16·1) |
| High-MDR/RR-TB burden^a^ | 5·41 (4·91–5·93) | 25·7 (23·4–28·0) | 2·87 (2·60–3·15) | 17·9 (16·4–19·4) | 2·29 (2·07–2·52) | 15·3 (14·0–16·6) |
|  | Income level^b^ | | | | | |
| LIC | 1·04 (0·95–1·15) | 4·20 (3·83–4·59) | 0·60 (0·54–0·66) | 3·31 (3·02–3·62) | 0·49 (0·44–0·55) | 2·97 (2·70–3·25) |
| LMIC | 5·12 (4·62–5·62) | 24·9 (22·6–27·1) | 2·71 (2·44–2·99) | 17·3 (15·8–18·8) | 2·16 (1·95–2·38) | 14·8 (13·6–16·1) |
| UMIC | 0·80 (0·72–0·88) | 3·21 (2·90–3·53) | 0·44 (0·39–0·48) | 2·25 (2·01–2·50) | 0·35 (0·32–0·39) | 1·94 (1·73–2·17) |
|  | World region | | | | | |
| AFR | 2·62 (2·41–2·86) | 11·3 (10·5–12·1) | 1·44 (1·32–1·58) | 8·52 (7·93–9·16) | 1·17 (1·06–1·28) | 7·49 (6·96–8·06) |
| AMR | 0·12 (0·11–0·13) | 0·41 (0·38–0·44) | 0·07 (0·06–0·07) | 0·30 (0·28–0·32) | 0·06 (0·05–0·06) | 0·26 (0·24–0·28) |
| EMR | 0·87 (0·73–1·03) | 3·11 (2·52–3·77) | 0·51 (0·42–0·60) | 2·35 (1·92–2·83) | 0·42 (0·35–0·49) | 2·08 (1·70–2·48) |
| EUR | 0·10 (0·09–0·11) | 0·22 (0·20–0·24) | 0·06 (0·05–0·07) | 0·15 (0·13–0·17) | 0·05 (0·04–0·06) | 0·13 (0·11–0·15) |
| SEAR | 2·40 (2·02–2·80) | 13·3 (11·4–15·3) | 1·22 (1·02–1·42) | 8·94 (7·68–10·3) | 0·96 (0·80–1·12) | 7·54 (6·48–8·64) |
| WPR | 0·85 (0·75–0·96) | 3·92 (3·42–4·52) | 0·45 (0·40–0·51) | 2·63 (2·29–3·04) | 0·36 (0·32–0·41) | 2·23 (1·93–2·57) |

Note: Values in parentheses represent equal-tailed 95% credible intervals. Catastrophic costs are defined as instances where the patient costs (either direct medical only or total) incurred during an episode of TB disease exceed either 10%, 20%, or 25% of total annual household income.

^a^ High-TB, high-TB/HIV (HIV-associated TB), and high-MDR/RR-TB (multidrug/rifampicin-resistant TB) burden countries as defined by the World Health Organization.

^b^ LIC: Gross national income (GNI) per capita of $1,085 or less; LMIC: GNI per capita of $1,086 to $4,225; UMIC: GNI per capita of $4,256 to $13,205 (World Bank 2021).

Note: All countries include 105 low- and middle-income countries analysed. AFR = African region; AMR = Region of the Americas; EMR = Eastern Mediterranean region; EUR = European region; GDP = gross domestic product; LIC = low-income; LMIC = lower middle-income; SEAR = Southeast Asian region; UMIC = upper middle-income; WPR = Western Pacific region.

**Appendix S19. Costs borne by TB-affected households averted by infant tuberculosis vaccines (in millions), costs discounted at 3%.**

| Country grouping | Direct medical costs borne by TB-affected households averted | Direct non-medical costs borne by TB-affected households averted | Indirect costs borne by TB-affected households averted | Total costs borne by TB-affected households averted |
| --- | --- | --- | --- | --- |
| All countries | 629 (560–693) | 1371 (1228–1516) | 1584 (1429–1757) | 3584 (3241–3953) |
| High-TB burden^a^ | 495 (431–554) | 1206 (1069–1353) | 1284 (1141–1442) | 2986 (2654–3332) |
| High-TB/HIV burden^a^ | 405 (343–461) | 1045 (910–1187) | 1123 (987–1273) | 2573 (2246–2910) |
| High-MDR/RR-TB burden^a^ | 477 (414–535) | 1081 (948–1223) | 1165 (1021–1322) | 2722 (2399–3068) |
|  | Income level^b^ | | | |
| LIC | 109 (87·4–132) | 190 (163–221) | 360 (293–434) | 659 (544–780) |
| LMIC | 422 (362–482) | 1045 (911–1191) | 940 (822·8–1073) | 2407 (2101–2748) |
| UMIC | 98·0 (84·4–112) | 136 (110–164) | 284 (210–372) | 518 (410–644) |
|  | World region | | | |
| AFR | 293 (242–338) | 654 (559–744) | 754 (644–866) | 1701 (1463–1929) |
| AMR | 16·9 (14·6–19·3) | 16·2 (14·5–18·0) | 17·6 (15·7–19·7) | 50·7 (45·5–56·1) |
| EMR | 115 (90·2–142) | 184 (141–230) | 311 (238–381) | 611 (474–752) |
| EUR | 13·2 (10·7–16·0) | 6·46 (5·53–7·37) | 9·03 (7·55–10·5) | 28·7 (23·8–33·6) |
| SEAR | 114 (86·3–149) | 376 (292–476) | 307 (242–390) | 797 (616–1018) |
| WPR | 76·3 (64·3–89·8) | 134 (107–162) | 185 (143–230) | 395 (317–477) |

Note: Values in parentheses represent equal-tailed 95% credible intervals. Total costs included patient direct medical, direct non-medical, and indirect costs (all undiscounted) in 2020 USD.

^a^ High-TB, high-TB/HIV (HIV-associated TB), and high-MDR/RR-TB (multidrug/rifampicin-resistant TB) burden countries as defined by the World Health Organization.

^b^ LIC: Gross national income (GNI) per capita of $1,085 or less; LMIC: GNI per capita of $1,086 to $4,225; UMIC: GNI per capita of $4,256 to $13,205 (World Bank 2021).

Note: All countries include 105 low- and middle-income countries analysed. AFR = African region; AMR = Region of the Americas; EMR = Eastern Mediterranean region; EUR = European region; GDP = gross domestic product; LIC = low-income; LMIC = lower middle-income; SEAR = Southeast Asian region; UMIC = upper middle-income; WPR = Western Pacific region.

**Appendix S20. Costs borne by TB-affected households averted by adolescent/adult tuberculosis vaccines (in millions), costs discounted at 3%.**

| Country grouping | Direct medical costs borne by TB-affected households averted | Direct non-medical costs borne by TB-affected households averted | Indirect costs borne by TB-affected households averted | Total costs borne by TB-affected households averted |
| --- | --- | --- | --- | --- |
| All countries | 4963 (4625–5302) | 10110 (9471–10810) | 11279 (10563–12109) | 26352 (24817–28081) |
| High-TB burden^a^ | 3798 (3496–4093) | 8700 (8062–9359) | 9332 (8659–10084) | 21831 (20374–23449) |
| High-TB/HIV burden^a^ | 2558 (2326–2806) | 7179 (6568–7825) | 7789 (7130–8517) | 17526 (16096–19046) |
| High-MDR/RR-TB burden^a^ | 3864 (3564–4190) | 7892 (7271–8533) | 8598 (7923–9334) | 20354 (18855–21937) |
|  | Income level^b^ | | | |
| LIC | 574 (504–650) | 1103 (1005–1208) | 1860 (1640–2101) | 3537 (3154–3934) |
| LMIC | 2742 (2484–3008) | 7044 (6421–7672) | 6280 (5743–6848) | 16067 (14699–17426) |
| UMIC | 1647 (1478–1829) | 1962 (1813–2122) | 3139 (2748–3547) | 6748 (6152–7345) |
|  | World region | | | |
| AFR | 1415 (1286–1560) | 3393 (3122–3678) | 4461 (4032–4928) | 9268 (8511–10094) |
| AMR | 298 (267–332) | 385 (355–415) | 372 (342–404) | 1056 (978–1133) |
| EMR | 567 (482–660) | 950 (793–1110) | 1448 (1212–1704) | 2965 (2516–3453) |
| EUR | 292 (248–343) | 155 (139–171) | 208 (182–235) | 655 (577–741) |
| SEAR | 1116 (936–1314) | 3714 (3192–4282) | 2879 (2449–3335) | 7709 (6586–8901) |
| WPR | 1275 (1128–1432) | 1513 (1372–1666) | 1911 (1692–2154) | 4699 (4249–5192) |

Note: Values in parentheses represent equal-tailed 95% credible intervals. Total costs included patient direct medical, direct non-medical, and indirect costs (all undiscounted) in 2020 USD.

^a^ High-TB, high-TB/HIV (HIV-associated TB), and high-MDR/RR-TB (multidrug/rifampicin-resistant TB) burden countries as defined by the World Health Organization.

^b^ LIC: Gross national income (GNI) per capita of $1,085 or less; LMIC: GNI per capita of $1,086 to $4,225; UMIC: GNI per capita of $4,256 to $13,205 (World Bank 2021).

Note: All countries include 105 low- and middle-income countries analysed. AFR = African region; AMR = Region of the Americas; EMR = Eastern Mediterranean region; EUR = European region; GDP = gross domestic product; LIC = low-income; LMIC = lower middle-income; SEAR = Southeast Asian region; UMIC = upper middle-income; WPR = Western Pacific region.
